# Effects of Ginkgo diterpene lactone meglumine on cognition and neurological function in stroke patients: a systematic evaluation and meta-analysis

**DOI:** 10.1101/2025.07.05.25330942

**Authors:** Fu-li Qin, Xia He, Xia-lian Huang, Yan-qiu Wang, Feng-le Mao, Yue-ming Cheng, Xiao-xue Zeng, Ming-xi Xu, Ying-ying Yang

## Abstract

**Objectives:** Evaluating the effect of GDLM on cognitive function and neurological prognosis in stroke patients

**Methods:** Investigators systematically searched nine databases for all randomized controlled trials (RCTs) from the inception of the databases to February 2025. Two investigators independently conducted literature screening, data extraction, bias risk assessment, and analyzed the data using data analysis software (StataMP 18).

**Results:** We included 34 RCTs involving 3,641 patients. Compared with the control group, the GDLM group showed significantly greater improvements in Montreal Cognitive Assessment (MoCA) (weighted mean difference [WMD] = 3.05, 95% confidence interval [CI] = 1.21 to 4.90), Mini-Mental State Examination (MMSE) (WMD = 3.68, 95% CI = 2.31 to 5.06), Barthel Index (BI) (WMD = 9.47, 95% CI = 8.02 to 10.91), and Activities of Daily Living Scale (ADLs) (WMD = 8.80, 95% CI = 6.59 to 11.00) scores. Additionally, more pronounced reductions were observed in National Institutes of Health Stroke Scale (NIHSS) (WMD = −2.30, 95% CI = −2.80 to −1.80) and modified Rankin Scale (mRS) (WMD = −0.45, 95% CI = −0.67 to −0.23) scores (all P < 0.001).

**Conclusion:** GDLM improves cognitive and neurological functions and enhances activities of daily living in stroke patients, supporting its clinical application. Future studies should involve longer follow-up, larger samples, and more rigorous RCTs to further clarify its effects on stroke patients.

## 1. Introduction

Stroke is a common cerebrovascular condition that develops clinically, caused by a sudden reduction in the supply of blood to brain cells, leading to ischemic damage and oxygen deprivation and resulting in structural and functional changes in the brain, affecting corresponding areas and resulting in roughly 87 percent of all cerebrovascular diseases (Feigin et al., 2022). Ischemic stroke and hemorrhagic stroke are the two basic classifications that are included in this illness (Barthels & Das, 2020; Carlew et al., 2021). Stroke can induce energy depletion, excessive glutamate release, free radical generation, and inflammatory responses, ultimately leading to a series of pathological events (Hankey, 2017), including widespread excitotoxic cell death. This disrupts the function of neural networks, resulting in sequelae such as sensory dysfunction and cognitive impairment (Durukan & Tatlisumak, 2007), which severely diminish patients’ quality of life (Coleman et al., 2017). Stroke is one of the major causes of disability worldwide and a significant contributor to neurological impairment and mortality among the elderly (Badawi et al., 2023). Additionally, Cognitive disability after stroke is a prevalent consequence (J.-H. Sun et al., 2014). It not only substantially increases the likelihood of stroke recurrence (Qu et al., 2015) but also greatly impedes the overall functional recovery of patients (Yuan et al., 2021) and is correlated with a notable rise in post-stroke mortality (Das et al., 2013). Research indicates that roughly one-third of stroke survivors subsequently acquire varied degrees of dementia (H. Kim et al., 2020; J.-H. Sun et al., 2014; Yuan et al., 2021). Therefore, the development of effective therapies targeting neuroprotection and cognitive recovery after stroke has become a research priority.

*Ginkgo biloba* L. is an ancient tree species that originated in China, and its leaves can treat diseases (Barker & Elston, 2022). In traditional Chinese medicine, *Ginkgo biloba* L. extract, derived from the leaves of the ginkgo tree, has long been used to treat cerebrovascular diseases (Tian et al., 2017; Q. Zhang et al., 2024). Growing evidence suggests their potential neuroprotective effects (Nabavi et al., 2015) and improved outcomes in ischemic stroke patients (S. Li et al., 2017). The Ginkgo diterpene lactone meglumine (GDLM) extracted from these leaves contains active components including ginkgolides A, B, and K (X. Chen et al., 2020). This patented Chinese herbal formulation (China drug approval number: Z20120024) has been certified by the National Medical Products Administration (NMPA) for treating mild-to-moderate cerebral infarction. Cerebral ischemia/reperfusion (I/R) injury after stroke dynamically induces a series of complex and dramatic metabolic perturbations. GDLM treatment can reduce I/R-induced brain injury by modulating energy metabolism, oxidative stress, and cerebral homeostasis, which are perturbed metabolic processes. In addition, the increased exposure of brain tissue to GDLM under pathological conditions may further enhance the neuroprotective effects of GDLM by directly targeting brain tissue (J.-L. Geng et al., 2017; T. Geng et al., 2018). At the same time, GDLM collectively protects against brain (I/R) injury by activating the Akt/Nrf2 pathway to mitigate oxidative stress injury, activating the Akt/CREB pathway to maintain neuronal survival, and inhibiting the TLR4/NF-κ B pathway activity to reduce pro-inflammatory factor release to attenuate inflammatory injury (S. Li et al., 2017; X. Li et al., 2020). Chen W et al (W. Chen et al., 2021). discovered that the active components in GDLM are a natural antagonist of platelet-activating factor (PAF) receptor, which effectively inhibits platelet aggregation, suppresses inflammatory response and prevents cell membrane damage. Based on the above and other extensive basic and clinical studies, GDLM reduces neuronal damage, protects neurological function, enhances cognitive function, and improves activities of daily living in stroke patients. With the rising number of RCTs, it has become an urgent requirement for meta-analyses to assess the efficiency of GDLM in treating cognitive dysfunction and neurological disability post-stroke. This meta-analysis evaluated the efficacy of GDLM in improving cognitive and neurological function in stroke survivors using standardized measures such as the Montreal Cognitive Assessment (MoCA), Mini-Mental State Examination (MMSE), National Institutes of Health Stroke Scale (NIHSS), modified Rankin Scale (mRS), Barthel Index (BI), and Activities of Daily Living Scale (ADLs). The results offer evidence-based guidance on clinical treatment.

## 2. Methods

The protocol was registered in PROSPERO on January 28, 2025 (CRD42025644270). Our study adhered to the Preferred Reporting Items for Systematic Review and Meta-Analyses 2020 statement (Page et al., 2021).

### 2.1. Search Strategy

Two investigators (Fu-li Q. & Yan-qiu W.) systematically searched databases and registries since their inception till February 2025, including PubMed, Embase, Cochrane Library, ClinicalTrials.gov, China National Knowledge Infrastructure (CNKI), Chinese Scientific Journal Database (VIP), SinoMed, Wanfang Database, and Chinese Clinical Trial Registry (ChiCTR). The search strategy combined English and Chinese keywords: (“Stroke” OR “Cerebral Stroke” OR “Cerebrovascular Accident”) AND (“ginkgo diterpene lactone meglumine injection” OR “ginkgo diterpene lactone meglumine” OR “ginkgo diterpene lactone”) (PubMed search strategy appears in Table S1).

### 2.2. Plant Nomenclature Validation

Plant species names extracted from the literature were standardized to currently accepted taxonomic names using The Plant List (www.theplantlist.org; accessed April 30, 2025), World Flora Online (www.worldfloraonline.org; accessed April 30, 2025), and Medicinal Plant Names Services (MPNS; http://mpns.kew.org; accessed April 30, 2025).

### 2.3. Inclusion and Exclusion Criteria

The inclusion criteria included: 1) Populations: Stroke patients; 2) Interventions: ginkgo diterpene lactone meglumine (GDLM); 3) Comparison: Control groups receiving standard care or placebo. Randomized controlled trials (RCTs), including those who experienced another conventional therapy (CT), were incorporated if the CT in the two groups was comparable; 4) The main results involved improvements in cognitive function and neurological status in stroke patients, evaluated with at least one of the following measurements: Montreal Cognitive Assessment (MoCA), Mini-Mental State Examination (MMSE), or National Institute of Health Stroke Scale (NIHSS). Secondary outcomes comprised the Modified Rankin Scale (mRS), Barthel Index (BI), and Activities of Daily Living Scale (ADLs); 5) Study design: RCTs.

The exclusion criteria encompassed: 1) Trials that do not include the desired outcomes; 2) Duplicate publications; 3) Self-controlled studies and nonRCTs; 4) Case reports, conference abstracts, preclinical studies, letters, conference abstracts, case analyses, review articles, systematic reviews and meta-analyses; 5) inability to obtain full text;

### 2.4. Study Selection

Literature selection was conducted independently by two investigators (Fu-li Q. & Yan-qiu W.). First, literature was imported into the reference manager EndNote 20, and duplicates were removed. Next, we reviewed the titles and abstracts of all publications against specified inclusion criteria and excluded those that were not. Finally, obtain and examine the whole texts for the remaining articles, thereafter, excluding those that fail to satisfy the inclusion criteria or fulfil any of the exclusion criteria. During this process, any disagreements were discussed and resolved; if necessary, a third investigator (Xia H.) engaged in the conversation to achieve unanimous agreement.

### 2.5. Data Extraction

Two investigators (Fu-li Q. & Yan-qiu W.) independently retrieved data from all qualifying trials, and any discrepancies were resolved through discussion with a professional investigator (Xia H.). The recorded study characteristics encompassed the following: (1) author(s); (2) publication year; (3) study design; (4) mean patient age; (5) sample size; (6) patient gender distribution; (7) disease duration; (8) intervention duration; (9) therapeutic protocols; (10) outcome measures assessing GDLM’s effects on stroke patients (MoCA, MMSE, NIHSS, mRS, BI, ADLs); (11) adverse events. When data was absent or not published in the research article, we emailed the corresponding authors to acquire complete information, if accessible.

### 2.6. Risk of bias assessment

Two independent evaluators (Fuli Q. & Yanqiu W.) assessed the risk of bias for the inclusion of RCTs using the Cochrane Risk of Bias Assessment Tool (ROB 2.0). The evaluation considered the following seven domains: (1) random sequence generation; (2) allocation concealment; (3) performance bias; (4) detection bias; (5) attrition bias; (6) reporting bias; and (7) other sources of bias. The following cases might lead to a bias risk assessment of “some concern” in the relevant domain: unclear allocation concealment despite baseline balance; potential awareness of the intervention by participants and investigators without confirmed deviations from the intended intervention, coupled with appropriate statistical methods to estimate intervention adherence effects; assessors’ awareness of the intervention with low likelihood of significantly affecting the study results. Disagreements were resolved through consultation with a third investigator (Xia H.) to reach a consensus.

### 2.7. Data Analysis

The meta-analysis was conducted using StataMP 18.0, incorporating assessments about potential publication bias, group-specific analyses, and sensitivity examinations. GDLM was employed as the intervention, with all outcome measures expressed as continuous variables and reported as mean ± standard deviation (SD). For outcomes measured with consistent units across studies, we calculated 95% confidence intervals (CI) and weighted mean differences (WMD), defined as the absolute difference between group means measured on identical scales. For outcomes with heterogeneous measurement units, standardized mean differences (SMD) were computed by dividing the between-group mean difference by the pooled standard deviation of participant outcomes, enabling data synthesis across trials employing different scales. A fixed-effect model was applied to outcomes with low heterogeneity (I² ≤ 50%), while a random-effects model was used for outcomes demonstrating substantial heterogeneity (I² > 50%).

### 2.8. subgroup analysis

Subgroups were analyzed according to stroke stage, intervention type, and treatment duration.

### 2.9. Sensitivity analysis

We employed a leave-one-out method to ascertain whether any individual study distorted the results, particularly for outcomes characterised by significant data variability. This method iteratively excluded each included study and recalculated the meta-analytic results to identify potential outliers disproportionately influencing heterogeneity or effect size magnitude.

### 2.10. publication bias

Publication bias was assessed using Egger’s regression test and funnel plots in StataMP 18.0 for meta-analyses involving ≥10 studies. Funnel plot symmetry was interpreted as indicating no publication bias; otherwise, potential bias was inferred.

## 3. Results

### 3.1. Studies included

Based on the searching methods, we were able to obtain 551 studies through the database. After deleting 218 duplicate studies with reference management software (EndNote 20), both the titles and the abstracts from the residual 333 papers were reviewed, resulting in the exclusion of 291 articles. The remaining 42 articles were thoroughly reviewed, leading to the exclusion of 8 articles for reasons including conference proceedings, incomplete data, and incomplete outcome measures. Ultimately, 34 articles (CAO Y. et al., 2021; CAO Z. et al., 2018; Chang, 2023; C. Chen et al., 2023; Dai et al., 2024; Feng et al., 2020; FENG et al., 2021; HAN et al., 2022; HU et al., 2021; HUANG, 2022; Jiang W., 2019; LI H. & ZHANG, 2021; LI Q. & MENG, 2023; LIU et al., 2019; MA et al., 2022; Miyasser & Bairkhaba, 2023; Qi et al., 2025; Sun et al., 2024; Sun Y. et al., 2019; J. Wang & Tan, 2024; Wang M. et al., 2016; WANG et al., 2020; Wang X. et al., 2017; XING et al., 2022; Yang G. et al., 2023; Yang Z. & Zhu, 2020; YIN & GONG, 2022; Zhang B. & Ye, 2020; M. Zhang et al., 2024; zhang et al., 2020; ZHANG et al., 2023; Zheng et al., 2023; ZHOU C. et al., 2024; ZHOU Y. et al., 2020) remained and were included in this study. (**Error! Reference source not found.**)

### 3.2. Study characteristics

The 34 included RCTs encompassed 3,641 stroke patients, including 1875 in the GDLM group and 1766 in the control group. (Table 1) Table 1 summarizes the baseline characteristics of the included studies, including region, study design, mean age, sex ratio, and intervention details. Among the 34 included RCTs, 28 trials (CAO Z. et al., 2018; C. Chen et al., 2023; Dai et al., 2024; Feng et al., 2020; FENG et al., 2021; HU et al., 2021; HUANG, 2022; Jiang W., 2019; LI H. & ZHANG, 2021; LI Q. & MENG, 2023; LIU et al., 2019; Miyasser & Bairkhaba, 2023; Qi et al., 2025; Sun Y. et al., 2019; J. Wang & Tan, 2024; Wang M. et al., 2016; WANG et al., 2020; Wang X. et al., 2017; XING et al., 2022; Yang G. et al., 2023; Yang Z. & Zhu, 2020; Zhang B. & Ye, 2020; M. Zhang et al., 2024; zhang et al., 2020; ZHANG et al., 2023; Zheng et al., 2023; ZHOU C. et al., 2024; ZHOU Y. et al., 2020) were conducted in the acute phase of stroke and 6 trials (CAO Y. et al., 2021; Chang, 2023; HAN et al., 2022; MA et al., 2022; Sun et al., 2024; YIN & GONG, 2022) in the recovery phase. All studies reported treatment duration, which ranged from 14 to 98 days. Specifically, 27 (CAO Y. et al., 2021; CAO Z. et al., 2018; C. Chen et al., 2023; Dai et al., 2024; Feng et al., 2020; FENG et al., 2021; HAN et al., 2022; HUANG, 2022; Jiang W., 2019; LI Q. & MENG, 2023; LIU et al., 2019; Miyasser & Bairkhaba, 2023; Qi et al., 2025; Sun Y. et al., 2019; J. Wang & Tan, 2024; Wang M. et al., 2016; WANG et al., 2020; Wang X. et al., 2017; Yang G. et al., 2023; YIN & GONG, 2022; Zhang B. & Ye, 2020; M. Zhang et al., 2024; zhang et al., 2020; ZHANG et al., 2023; Zheng et al., 2023; ZHOU C. et al., 2024; ZHOU Y. et al., 2020) trials had a 14-day duration, 4 trials (HU et al., 2021; LI H. & ZHANG, 2021; Sun et al., 2024; XING et al., 2022) lasted 28 days, 1 trial (Yang Z. & Zhu, 2020) lasted 8 weeks, 1 trial (Chang, 2023) lasted 12 weeks, and1 trial (MA et al., 2022) lasted 14 weeks. The interventions were classified into four categories: 8 trials (HU et al., 2021; LIU et al., 2019; Miyasser & Bairkhaba, 2023; WANG et al., 2020; Yang G. et al., 2023; Yang Z. & Zhu, 2020; zhang et al., 2020; ZHOU C. et al., 2024) assessed the effectiveness of GDLM combined with neuroprotective agents (e.g., Edaravone, Butylphthalide), 15 trials (CAO Y. et al., 2021; Chang, 2023; C. Chen et al., 2023; Dai et al., 2024; Feng et al., 2020; LI H. & ZHANG, 2021; LI Q. & MENG, 2023; MA et al., 2022; Qi et al., 2025; Sun Y. et al., 2019; J. Wang & Tan, 2024; XING et al., 2022; Zhang B. & Ye, 2020; M. Zhang et al., 2024; ZHANG et al., 2023) assessed GDLM combined with antiplatelet or anticoagulant medications (e.g., Aspirin, Clopidogrel, Tirofiban), 6 trials (CAO Y. et al., 2021; Chang, 2023; HAN et al., 2022; MA et al., 2022; Sun et al., 2024; YIN & GONG, 2022) assessed GDLM paired with neurotrophic/metabolic therapies (e.g., Troxerutin, Oxiracetam, and rehabilitation training), and 5 trials (CAO Z. et al., 2018; HUANG, 2022; Jiang W., 2019; M. Zhang et al., 2024; ZHOU Y. et al., 2020) assessed GDLM integrated with guideline-based conventional symptomatic therapies (interventions not detailed). Outcomes assessed included post-stroke cognitive function, neurological function, and activities of daily living. Specifically, 4 trials (Jiang W., 2019; Yang Z. & Zhu, 2020; YIN & GONG, 2022; M. Zhang et al., 2024) reported MOCA as an outcome measure, 9 trials (Chang, 2023; Dai et al., 2024; HAN et al., 2022; HUANG, 2022; Jiang W., 2019; MA et al., 2022; Miyasser & Bairkhaba, 2023; Yang G. et al., 2023; M. Zhang et al., 2024) reported MMSE, 32 trials (CAO Y. et al., 2021; CAO Z. et al., 2018; Chang, 2023; C. Chen et al., 2023; Dai et al., 2024; Feng et al., 2020; FENG et al., 2021; HAN et al., 2022; HU et al., 2021; HUANG, 2022; LI H. & ZHANG, 2021; LI Q. & MENG, 2023; LIU et al., 2019; Miyasser & Bairkhaba, 2023; Qi et al., 2025; Sun et al., 2024; Sun Y. et al., 2019; J. Wang & Tan, 2024; Wang M. et al., 2016; WANG et al., 2020; Wang X. et al., 2017; XING et al., 2022; Yang G. et al., 2023; Yang Z. & Zhu, 2020; YIN & GONG, 2022; Zhang B. & Ye, 2020; M. Zhang et al., 2024; zhang et al., 2020; ZHANG et al., 2023; Zheng et al., 2023; ZHOU C. et al., 2024; ZHOU Y. et al., 2020) reported NIHSS, 14 trials (CAO Z. et al., 2018; Chang, 2023; Dai et al., 2024; Feng et al., 2020; FENG et al., 2021; HAN et al., 2022; HU et al., 2021; LI H. & ZHANG, 2021; LI Q. & MENG, 2023; Qi et al., 2025; WANG et al., 2020; Zhang B. & Ye, 2020; zhang et al., 2020; ZHOU Y. et al., 2020) reported Barthel Index, 6 trials (HUANG, 2022; Sun et al., 2024; Wang M. et al., 2016; Yang G. et al., 2023; M. Zhang et al., 2024; ZHOU C. et al., 2024) reported ADLs, and 7 trials (C. Chen et al., 2023; Feng et al., 2020; Qi et al., 2025; Sun et al., 2024; Sun Y. et al., 2019; Wang X. et al., 2017; zhang et al., 2020) reported mRS as an outcome measures.

**Table 1.**
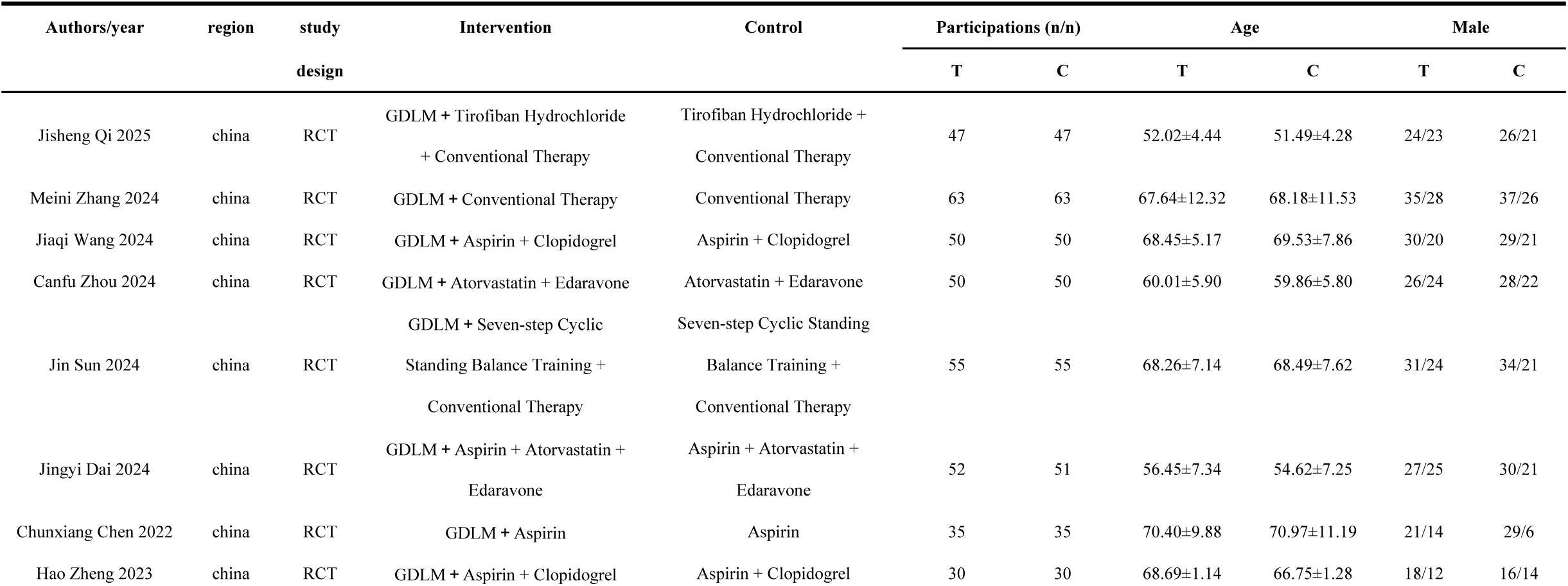
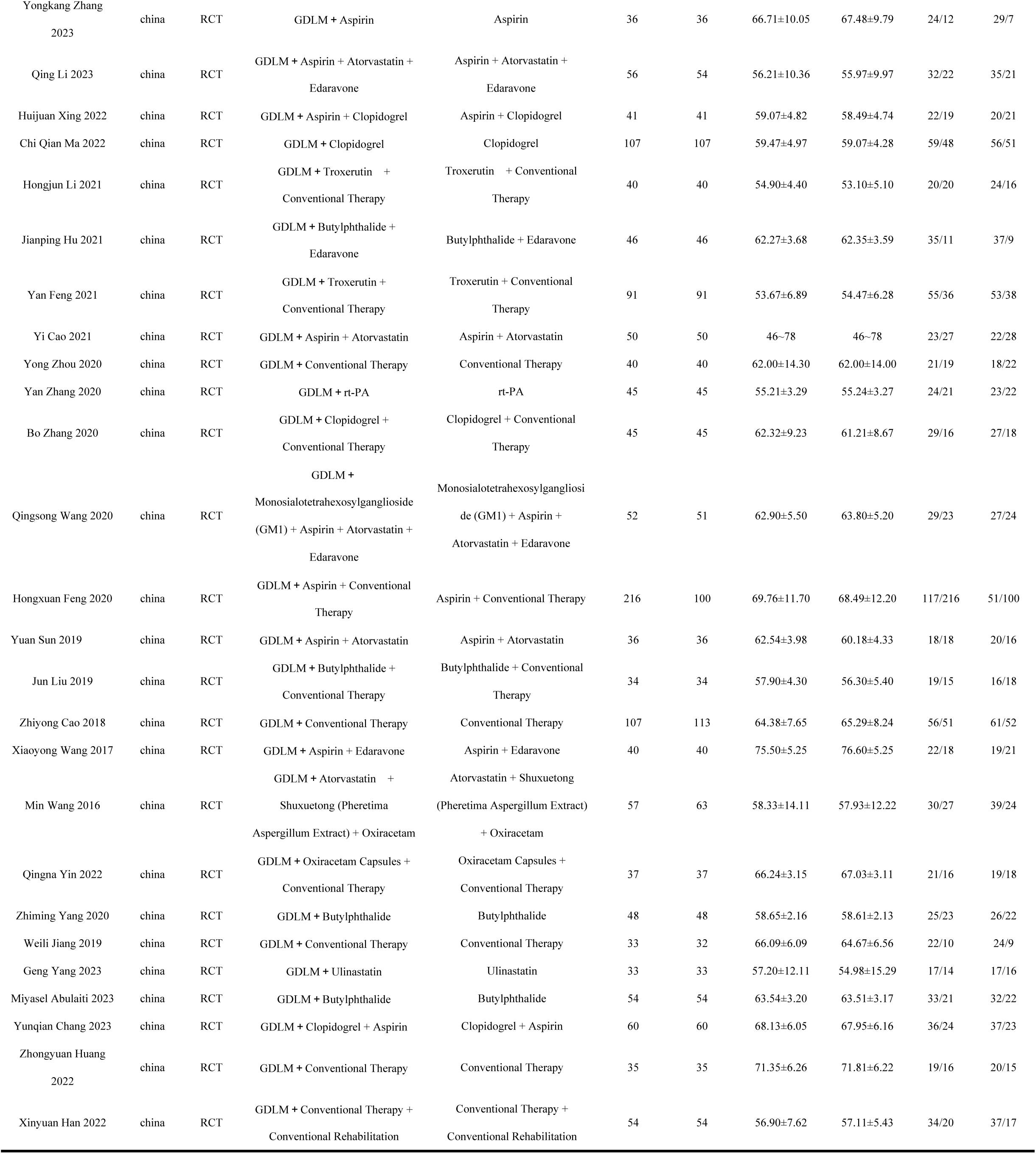
Basic characteristics of included trials.

### 3.3. Evaluation of the Quality of the Included Studies

Trials were categorized into three levels based on the number of components with a potentially high risk of bias (ROB): high risk, moderate risk, and low risk (Higgins et al., 2011). The 34 involved articles comprised RCTs, of which 4 were classified as low risk and 30 as moderate risk. The specific details are provided below (**Error! Reference source not found.**). (**Error! Reference source not found.**) illustrates the risk of bias assessment for the included studies.

### 3.4. Efficacy outcomes

#### 3.4.1. Montreal Cognitive Assessment ( MOCA )

Four (Jiang W., 2019; Yang Z. & Zhu, 2020; YIN & GONG, 2022; M. Zhang et al., 2024) RCTs reported using MoCA scores as an outcome index. About 361 patients participated, comprising 181 within the GDLM group and 180 in the control group. The analysis indicated a more pronounced increase in MOCA scores within the GDLM group relative to the control group, exhibiting a weighted mean difference of 3.05, accompanied by a 95% confidence interval of 1.21 to 4.90 (WMD = 3.05, 95% CI = 1.21–4.90, p < 0.001, I² = 69.42%). The two groups showed a substantial amount of heterogeneity (Heterogeneity: Tau² = 2.32, p = 0.01, I^2^ = 69.42 %). (**Error! Reference source not found.**)

#### 3.4.2. Mini-Mental State Examination ( MMSE )

Nine (Chang, 2023; Dai et al., 2024; HAN et al., 2022; HUANG, 2022; Jiang W., 2019; MA et al., 2022; Miyasser & Bairkhaba, 2023; Yang G. et al., 2023; M. Zhang et al., 2024) RCTs comparing MMSE scores in the GDLM group and the control group revealed significant heterogeneity between the studies (Heterogeneity: Tau² = 3.16, p<0.001, I^2^ = 81.8 %). A total of 980 patients were enrolled in the nine RCT trials, including 491 in the GDLM group and 489 in the control group. A meta-analysis utilizing a random-effects model indicated that the increase in MMSE scores had been much greater in the GDLM group compared to the control group (WMD = 3.68, 95% CI = 2.31-5.06, p < 0.001). (**Error! Reference source not found.**)

#### 3.4.3. National Institute of Health Stroke Scale ( NIHSS )

This analysis included 32 RCTs (CAO Y. et al., 2021; CAO Z. et al., 2018; Chang, 2023; C. Chen et al., 2023; Dai et al., 2024; Feng et al., 2020; FENG et al., 2021; HAN et al., 2022; HU et al., 2021; HUANG, 2022; LI H. & ZHANG, 2021; LI Q. & MENG, 2023; LIU et al., 2019; Miyasser & Bairkhaba, 2023; Qi et al., 2025; Sun et al., 2024; Sun Y. et al., 2019; J. Wang & Tan, 2024; Wang M. et al., 2016; WANG et al., 2020; Wang X. et al., 2017; XING et al., 2022; Yang G. et al., 2023; Yang Z. & Zhu, 2020; YIN & GONG, 2022; Zhang B. & Ye, 2020; M. Zhang et al., 2024; zhang et al., 2020; ZHANG et al., 2023; Zheng et al., 2023; ZHOU C. et al., 2024; ZHOU Y. et al., 2020), all of which used the NIHSS scale for efficacy evaluation, with an aggregate sample size of 3362 patients, of which 1735 were in the GDLM group while 1627 were in the control group. Meta-analysis found a more pronounced decrease in NIHSS scores within the GDLM group relative to the control group, with a weighted mean difference in NIHSS scores of −2.30, 95% confidence interval of −2.80 to −1.80 (WMD = −2.30, 95% CI = −2.80 - −1.80, p<0.001). The results showed high heterogeneity (Heterogeneity: Tau^2^ = 1.24, p<0.001, *I*² = 82.31%). (**Error! Reference source not found.**)

#### 3.4.4. Barthel Index ( BI )

Fourteen (CAO Z. et al., 2018; Chang, 2023; Dai et al., 2024; Feng et al., 2020; FENG et al., 2021; HAN et al., 2022; HU et al., 2021; LI H. & ZHANG, 2021; LI Q. & MENG, 2023; Qi et al., 2025; WANG et al., 2020; Zhang B. & Ye, 2020; zhang et al., 2020; ZHOU Y. et al., 2020) RCTs compared BI scores in the GDLM and control groups. An aggregate of 1788 participants were recruited, of whom 951 were assigned to the GDLM group and 837 to the control group. Meta-analysis showed a significantly higher increase in BI scores in the GDLM group than in the control group, with a weighted mean difference in BI scores of 9.47, 95% confidence interval of 8.02 to 10.91 (WMD =9.47,95 % CI = 8.02-10.91, p < 0.001). The results showed low heterogeneity (Heterogeneity:H^2^ = 1.65, p < 0.001, I^2^ = 39.26%). (**Error! Reference source not found.**)

#### 3.4.5. Activities of Daily Living Scale (ADLs)

Six RCTs (HUANG, 2022; Sun et al., 2024; Wang M. et al., 2016; Yang G. et al., 2023; M. Zhang et al., 2024; ZHOU C. et al., 2024) involving 592 participants assessed ADLs between 293 patients in the GDLM group and 299 patients in the control group. According to the meta-analysis, the increase in ADLs scores was significantly higher in the GDLM group than in the control group, while the weighted mean difference in ADLs was 8.80, with a 95 % confidence interval of 6.59 to 11.00 (WMD = 8.80, 95 % CI = 6.59 to 11.00, p < 0.001). The results showed moderate heterogeneity (Heterogeneity: Tau^2^ =3.46, p< 0.001, *I*² = 54.13%). (**Error! Reference source not found.**)

#### 3.4.6. Modified Rankin Scale ( mRS )

Seven RCTs (C. Chen et al., 2023; Feng et al., 2020; Qi et al., 2025; Sun et al., 2024; Sun Y. et al., 2019; Wang X. et al., 2017; zhang et al., 2020) with 474 patients in the GDLM group and 358 patients in the control group utilized the mRS assessment results as outcome indicators. Pooled analysis showed a more pronounced reduction in mRS scores in the GDLM group relative to the control group (WMD = −0.45, 95% CI = - 0.67 to −0.23), with barely any heterogeneity observed between the two groups (Heterogeneity: H² = 1.00, p < 0.001, I² = 0.00%). (**Error! Reference source not found.**)

### 3.5. Subgroup analysis

Subgroup analyses were conducted across three ways: stroke phase, intervention methods and treatment duration (**Error! Reference source not found.**). Subgroup analysis based on stroke stage showed that the difference was not statistically significant in the reduction of mRS scores within the GDLM group and the control group in patients at the recovery stage (P>0.05). But a substantial distinction was found in patients within the acute phase (P < 0.001). In the subgroup analysis of intervention methods, when the control group received neuroprotective agents or neurotrophic/metabolic improvement therapies as conventional interventions, the difference in mRS score reduction within the GDLM group as well as the control group was not statistically significant (P>0.05). Similarly, in studies where the control group received standard routine treatment, the increases in MOCA and MMSE scores in the GDLM treatment group compared with the control group were not statistically significant (P>0.05). Meanwhile, the subgroup analysis of treatment duration indicated that the 28-day intervention failed to result in a statistically significant reduction in the mRS score for the GDLM group relative to the control group (P>0.05).

### 3.6. Sensitivity analysis

In our study, to evaluate the impact of every single study on the overall WMD, we conducted a leave-one-out analysis on the MoCA, MMSE, NIHSS, BI, ADLs, and mRS scores among the GDLM group as well as the control group. The results of the sensitivity analysis showed that regardless of which study was excluded, the combined WMD for MoCA, MMSE, NIHSS, BI, ADLs, and mRS scores remained unchanged within the GDLM group and the control group. (**Error! Reference source not found.**)

### 3.7. publication bias

We assessed the presence of publication bias in the findings from the NIHSS and BI scores. For NIHSS scores, Egger ’ s test (p = 0.0033) and the funnel plot both demonstrated the presence of publication bias. In contrast, for BI scores, both Egger’s test (p = 0.8499) and the funnel plot suggested the absence of publication bias. (**Error! Reference source not found.**)

## 4. Discussion

This meta-analysis was mainly intended to evaluate the efficacy of GDLM on cognitive ability, neurological function, and activities of daily living within stroke patients. Our analysis included 34 RCTs involving 3,641 patients diagnosed with stroke. The results of our analysis indicate that the GDLM group experienced more substantial increases in MOCA, MMSE, BI, and ADLs scores than the control group. Concurrently, the reductions in NIHSS and mRS scores appeared considerably greater in the GDLM group compared to the control group.

Stroke is the second largest cause of death worldwide (Donkor, 2018) and one of the most prevalent cerebrovascular illnesses (Feigin et al., 2022). Furthermore, it is distinguished by high rates of impairment, recurrence, and occurrence (M. Zhang et al., 2024). After a stroke, a sudden drop in cerebral perfusion triggers a decrease in local oxygen metabolism, and ischemia and hypoxia not only lead to cerebral circulatory disorders and an imbalance of energy metabolism but also activate local immune responses, resulting in a dynamic imbalance between pro-inflammatory factors (IL-1β, TNFα) and anti-inflammatory factors (IL-10, TGFβ) (J.-T. Kim et al., 2020; Orellana-Urzúa et al., 2020; Zhao et al., 2022). In addition, the aggregation of inflammatory cytokines not only leads to the disruption of the integrity of the blood-brain barrier (BBB) but also promotes the accumulation of oxygen-free radicals and the abnormal release of neurotransmitters, which ultimately induce neuronal damage and apoptosis, leading to neurological and cognitive deficits (Dl et al., 2023; Jurcau & Simion, 2021; C. Yang et al., 2019). The core problem in stroke is the loss of neuronal cells, which makes it difficult or even impossible for sub-acute patients to recover in the late stages (Fan et al., 2020). Therefore, therapies targeting neuronal cell damage and apoptosis are a critical focus in current clinical treatment. In recent years, there has been growing research interest in plant-derived extracts from traditional medicine for the treatment of stroke. GDLM is a pure traditional Chinese medicine preparation, primarily derived from the traditional Chinese herb *Ginkgo biloba* L. (Fan et al., 2020). Its main active components are the diterpene lactones of Ginkgo biloba, including ginkgolide A, B, and K. Previous studies have demonstrated that GDLM exerts multiple clinical benefits, including promoting blood circulation to remove blood stasis, unblocking the meridians to relieve pain, astringing the lungs to relieve asthma, resolving phlegm, and reducing blood lipids (T. Geng et al., 2018; Xiao et al., 2015). Initially, studies demonstrate that ginkgolide B, an active component in GDLM, is the most potent PAF antagonist. It can eliminate PAF-induced platelet aggregation, thereby inhibiting thrombus formation and alleviating neurological deficits in patients with cerebral infarction (CAO H. & ZHANG, 2019; Zhang H. et al., 2020). Subsequently, GDLM reduces BBB permeability, alleviating cerebral edema, improving cerebral metabolic dysregulation, and suppressing neuronal apoptosis (X. Li et al., 2020). Thereafter, GDLM treatment can increase the serum levels of neurotrophic factors Neurturin and IGFBP2, thereby exerting a neuroprotective effect on neuronal cells (Burgdorf et al., 2017). Ultimately, GDLM mitigates brain injury by suppressing the release of inflammatory factors (e.g., IL-6, IL-8), thereby attenuating inflammatory responses and oxidative stress while preserving neurological function (S. Jiang et al., 2021). P300, as a classic endogenous component of event-related potentials (ERPs), is widely used in clinical settings to assess the severity of cognitive impairment (H. Li et al., 2021). Wang et al (Wang X. et al., 2023). demonstrated that after intervention with GDLM, the latency of the P300 component was significantly shortened, while its amplitude was notably increased, indicating enhanced efficiency of neural information processing. Further research has revealed that GDLM may exert its effects on post-stroke cognitive function by modulating electroencephalographic responses to external stimuli, thereby mediating neuroplasticity effects related to cognitive deficits following stroke (M. Zhang et al., 2024). Additionally, GDLM has been proven to improve cognitive performance via several pathways. It lowers fibrinogen (FIB) and D-dimer (DD) levels, decreasing blood viscosity and improving hypercoagulability (M. Zhang et al., 2024). These effects enhance cerebral microcirculation, allowing for greater oxygen and nutrition delivery to the brain areas (Orellana-Urzúa et al., 2020). Furthermore, GDLM enhances the regeneration of injured neurons, which is critical for regaining cognitive functions after stroke. Collectively, GDLM exerts its therapeutic effects through multi-target synergistic actions, protecting neuronal cells, improving cerebral hemorheological parameters, reducing whole blood viscosity, alleviating neuronal apoptosis, and promoting neural repair. These mechanisms ultimately lead to significant improvements in cognitive function and neurological deficits in stroke patients, as well as enhanced activities of daily living.

Our subgroup analysis revealed no significant differences in mRS score improvement between GDLM and control groups among patients in the stroke recovery phase, those receiving neuroprotective agents or neurotrophic/metabolic improvement interventions, or those undergoing 4-week treatment courses (all P > 0.05). We posit that this distinction might be explained by an insufficient number of subgroup samples, notably with only one study included in each subgroup. Furthermore, in the study where the control group received standard conventional therapy, the MOCA and MMSE scores of the GDLM group did not show substantial enhancement when compared to the control group, which may be related to the specificity of the intervention mechanism and insufficient treatment duration. Standard conventional treatments (such as antihypertensive therapy, oxygen administration, and cerebral cell nutrition) are generally supportive measures targeting post-stroke conditions and have broad mechanisms of action that may partially overlap with the targets of GDLM (e.g., improving blood flow or providing neuroprotection), thereby masking the specific therapeutic effects of GDLM. In contrast, neuroprotective agents (such as Butylphthalide and Human Urinary Kallidinogenase [HUK]) and antiplatelet/anticoagulant therapies, which act through synergistic mechanisms (e.g., by inhibiting inflammation or improving cerebral microcirculation), can significantly enhance the efficacy of GDLM. Our study also has several limitations. First, though the literature search stage did not limit the language type, of the 34 articles finally included, 31 had been published in Chinese and just 3 in English, and no relevant research in other languages was located. Second, all RCTs were conducted in China, which may limit the generalizability of the study results. Additionally, although we performed sensitivity and subgroup analyses, we still did not identify the source of high heterogeneity in our study results, considering potential confounding factors, which reduces the reliability of our findings. Therefore, in future studies, we should expand the scope of the study population, conduct precise stratified RCTs based on stroke subtypes and biomarkers, and systematically explore the dose-time effect relationship to determine the optimal intervention window and the minimum effective duration of treatment. However, our study represents the most recent meta-analysis of the efficacy of GDLM on cognitive and neurological functions in stroke patients, offering evidence-based medical assistance for clinical practice.

## 5. Conclusion

Our meta-analysis demonstrates that GDLM has positive effects on the recovery of cognitive function, neurological function, and activities of daily living in patients after stroke. However, considering the study’s limitations, including regional selection bias, potential heterogeneity, and the impact of social support systems, subsequent clinical research should utilize high-quality RCTs with substantial numbers of samples and extended durations to further ascertain whether GDLM can enhance cognitive abilities, neurological function, and activities of daily living in stroke patients.

## Data Availability

All data underlying the findings of this meta-analysis are derived from previously published studies, which are fully cited in the References section.

## CRediT authorship contribution statement

Fu-li Qin: Writing – original draft, Visualization, Validation, Software, Investigation. Xia He: Writing – review & editing, Supervision, Conceptualization. Xia-lian Huang: Writing –review & editing, Supervision, Project administration, Funding acquisition, Conceptualization. Yan-qiu Wang: Methodology, Visualization, Investigation. Feng-le Mao: Methodology, Investigation. Yue-ming Cheng: Methodology, Investigation. Xiao-xue Zeng: Visualization, Investigation. Ming-xi Xu: Software, Investigation. Ying-ying Yang: Software, Investigation

## Availability of data and materials

Data sharing does not apply to this article, as no datasets were generated or analyzed during the current study

---

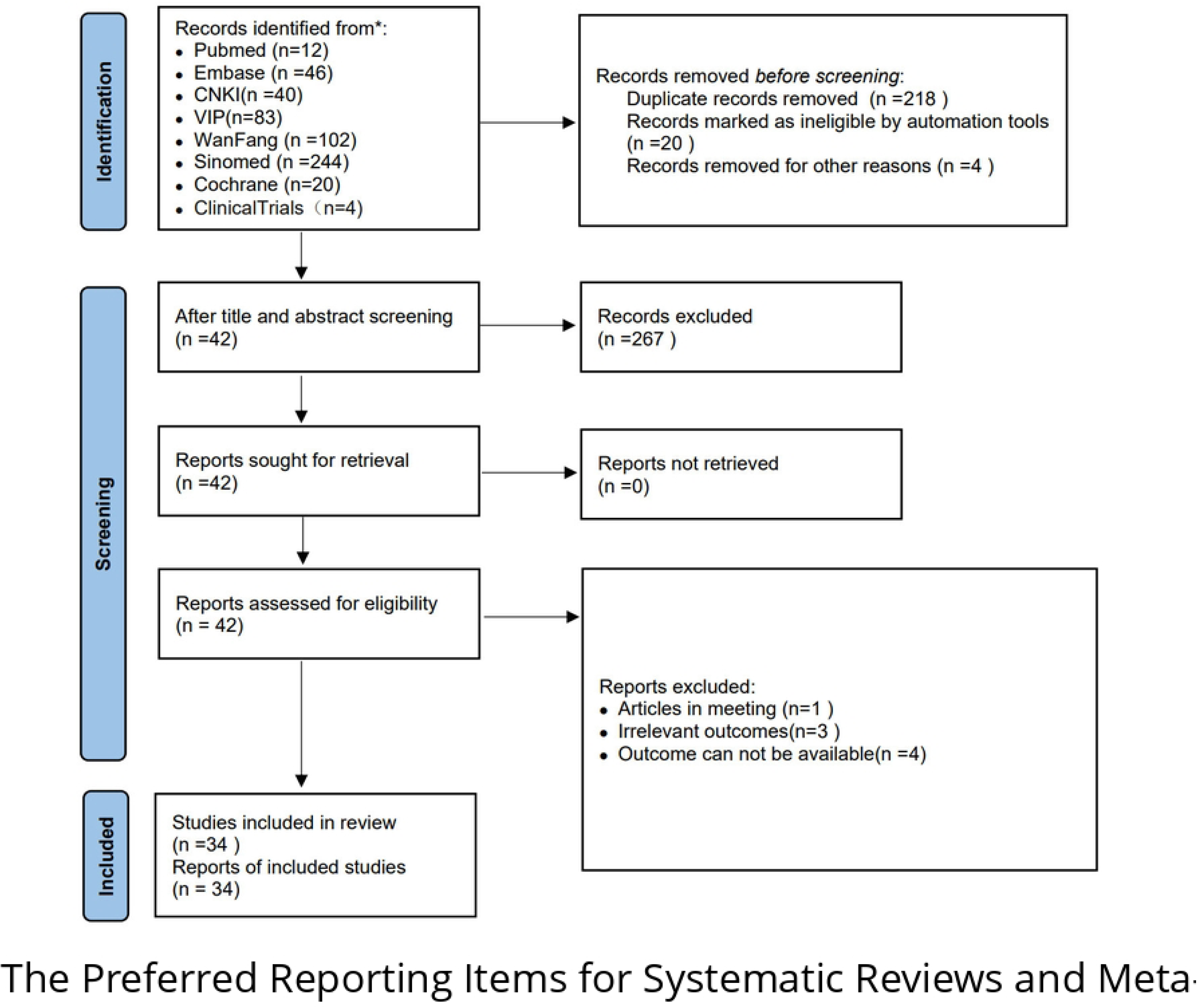

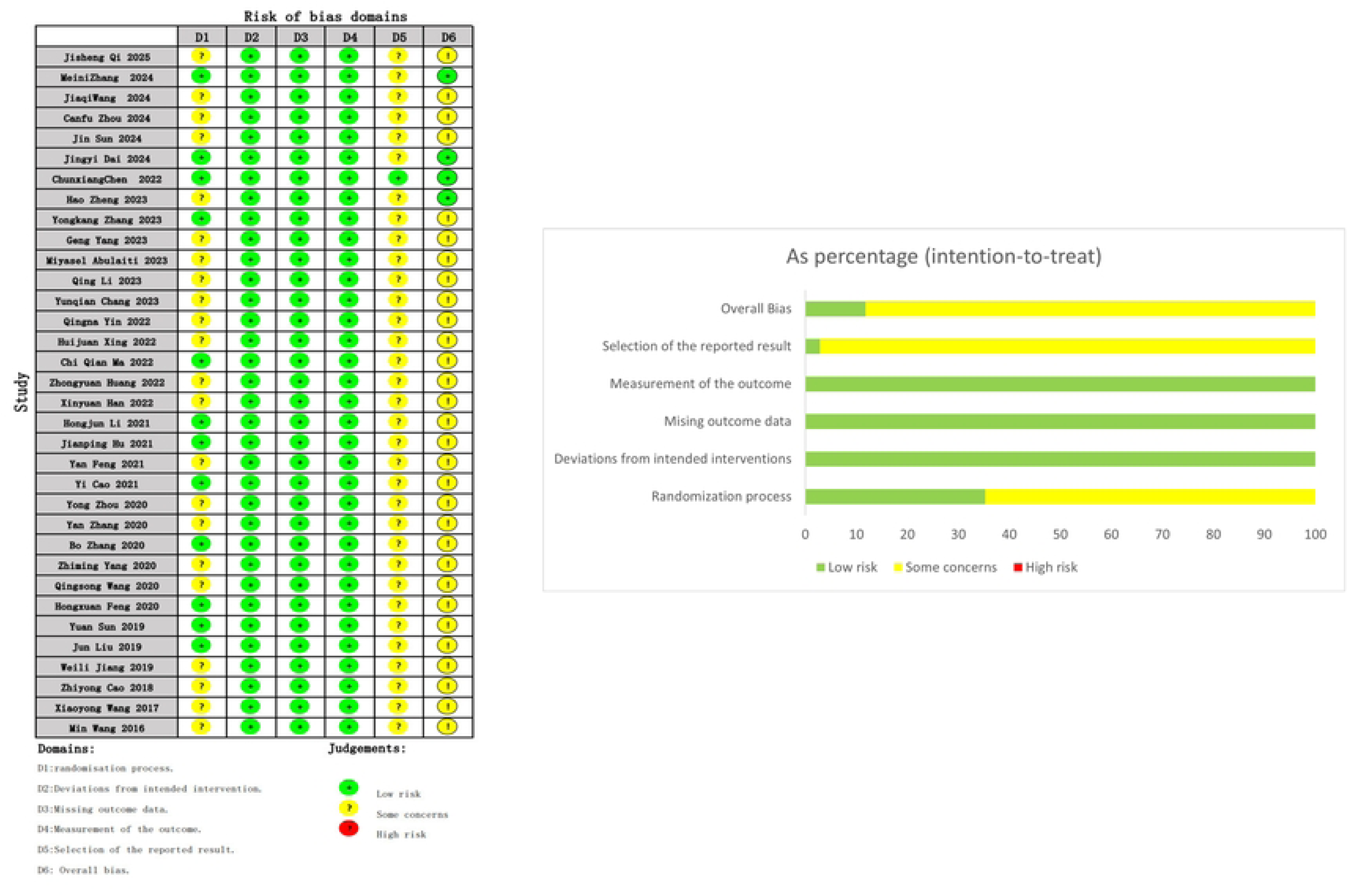

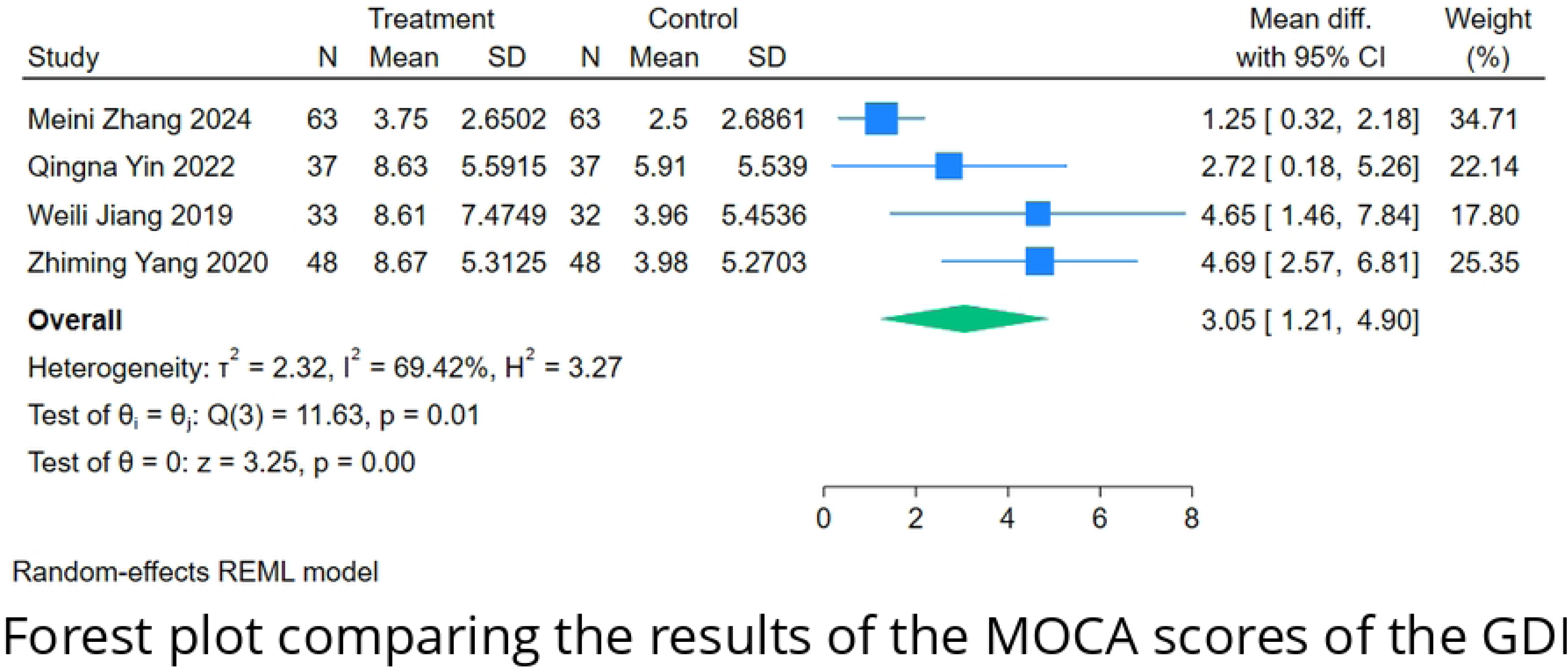

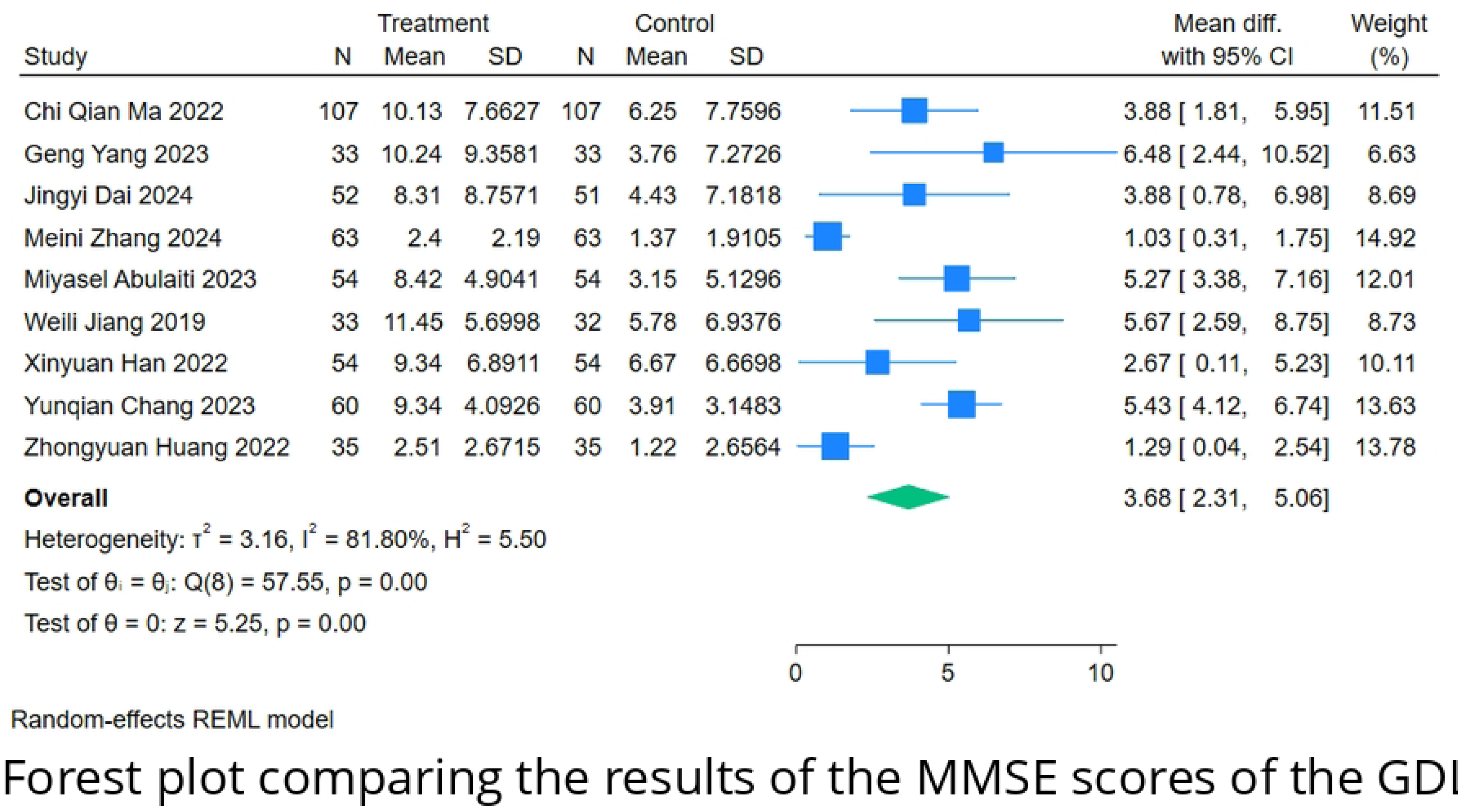

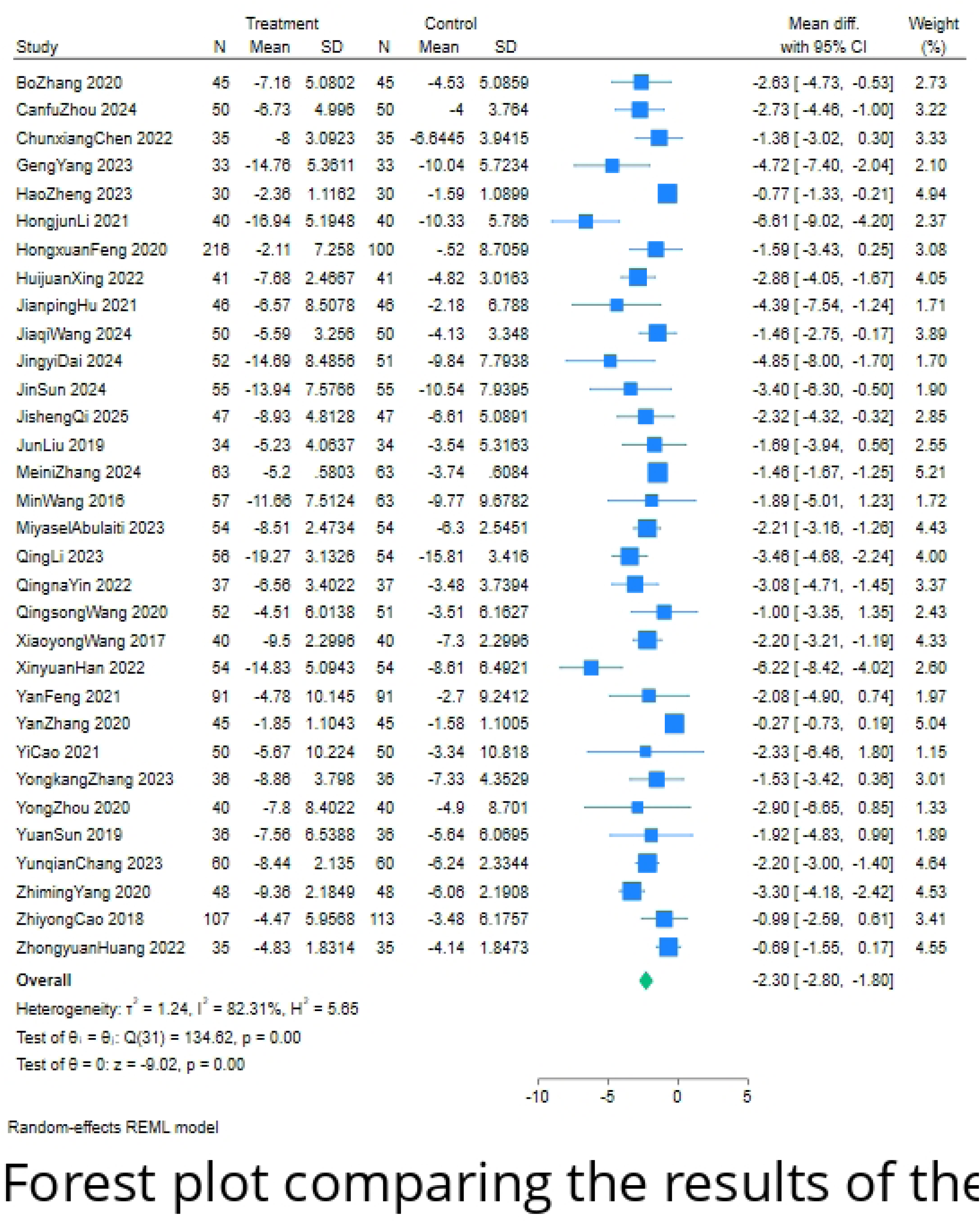

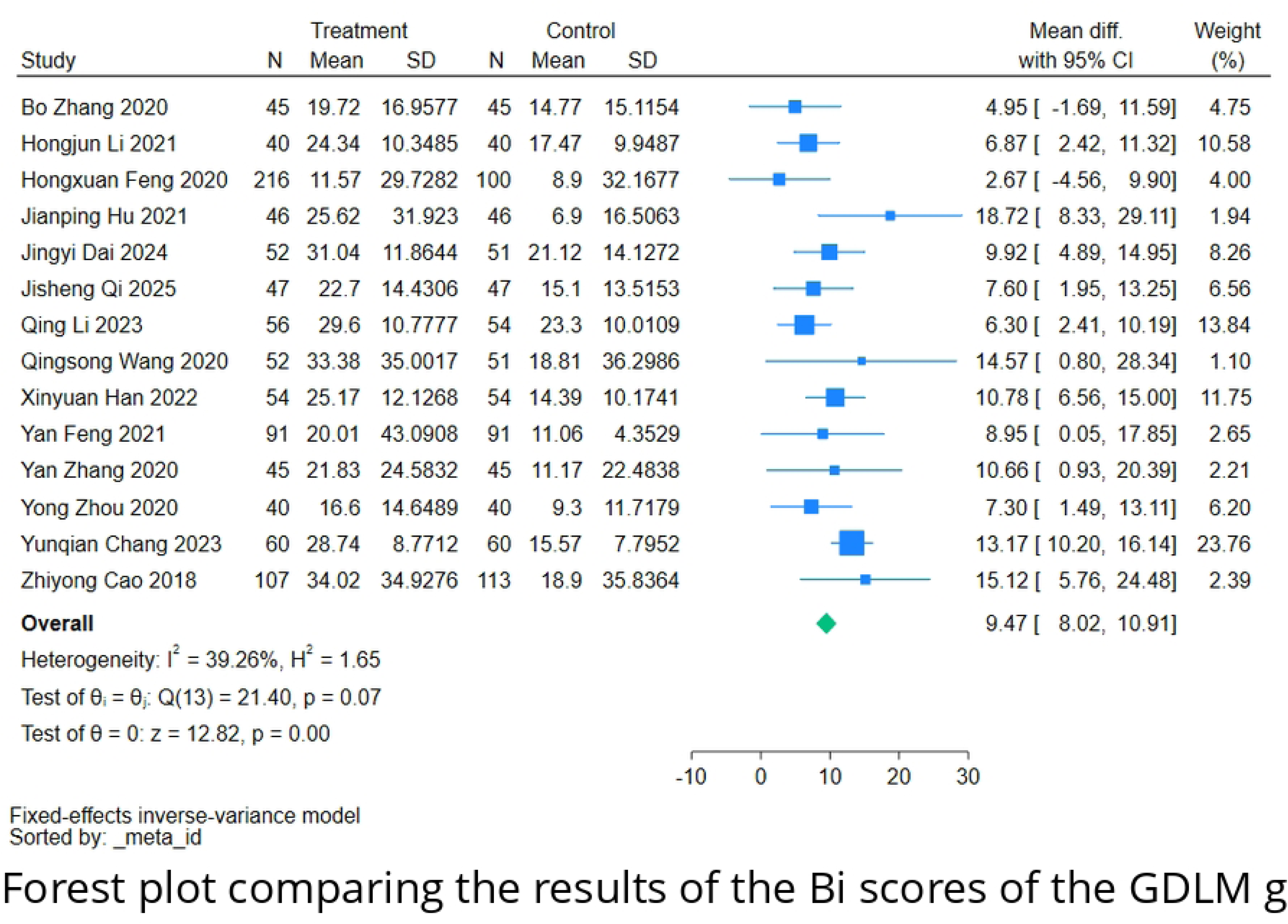

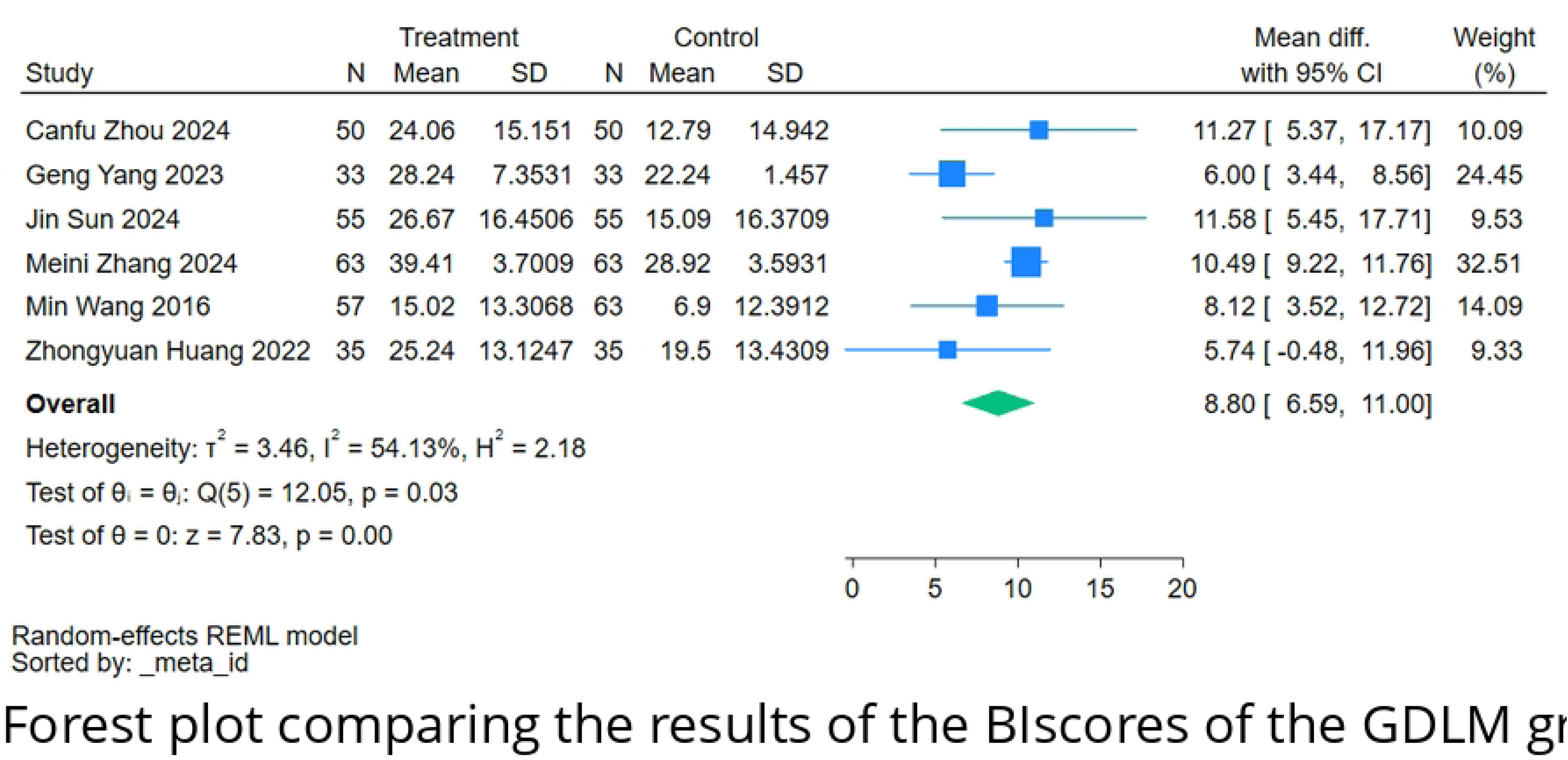

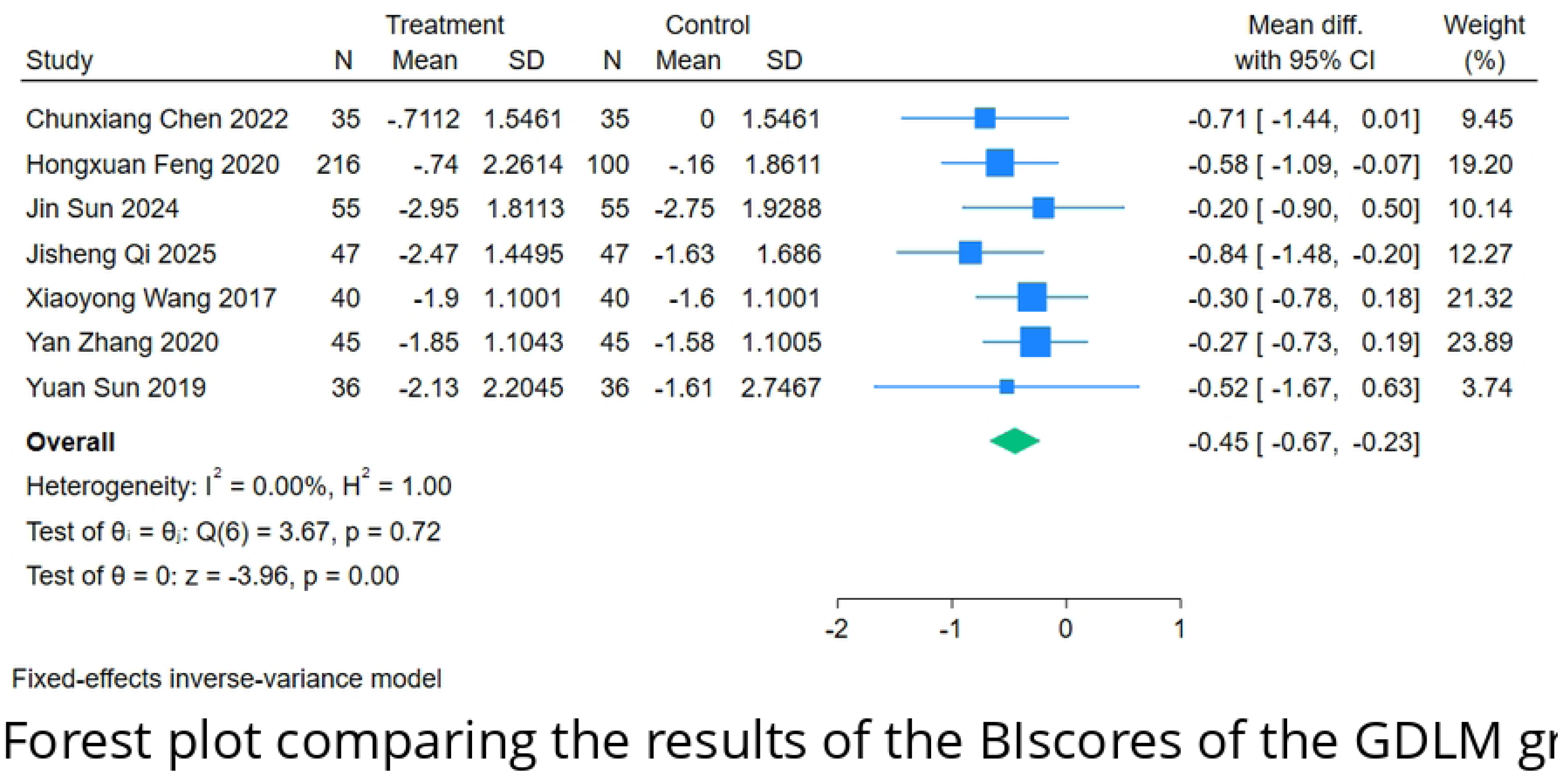

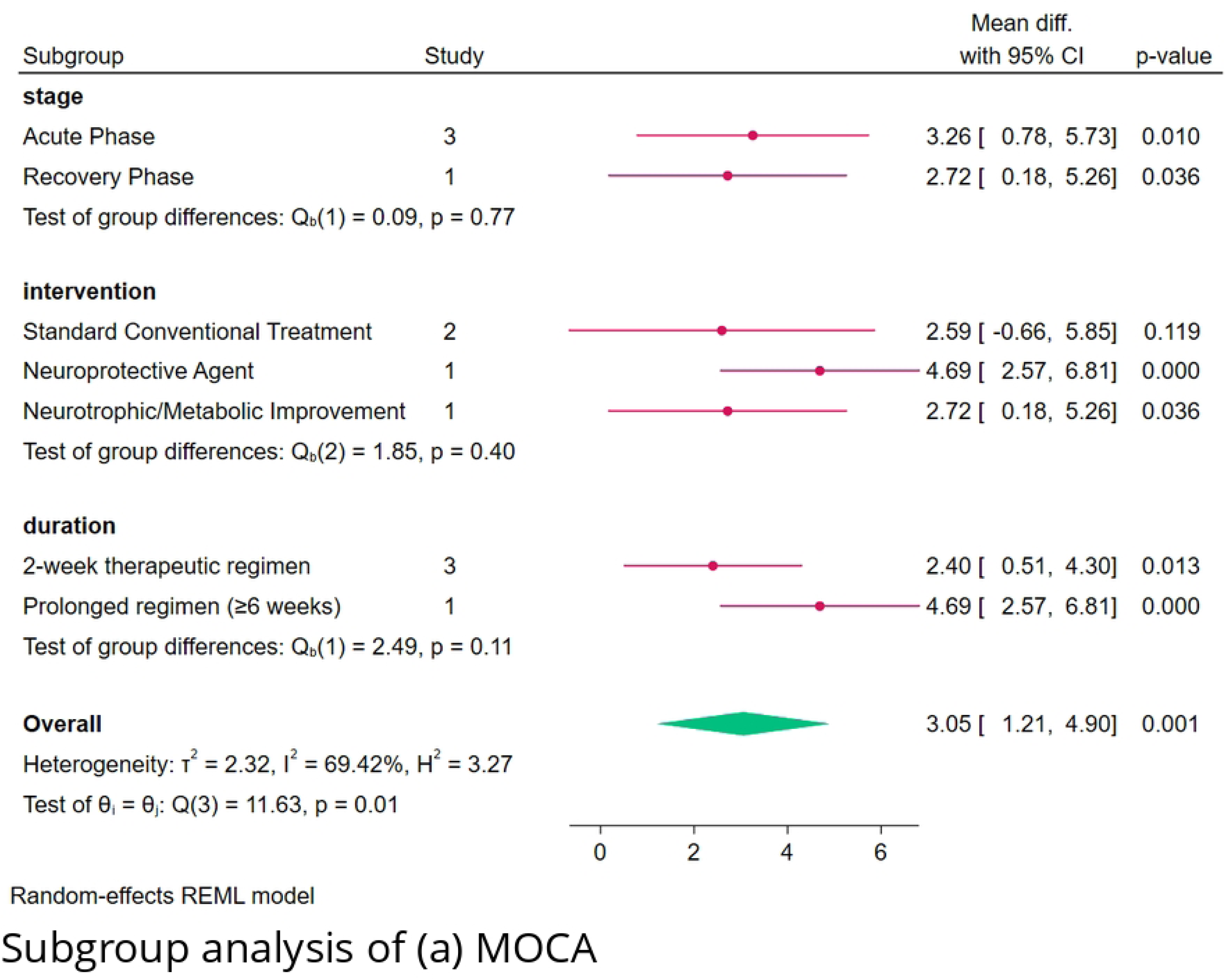
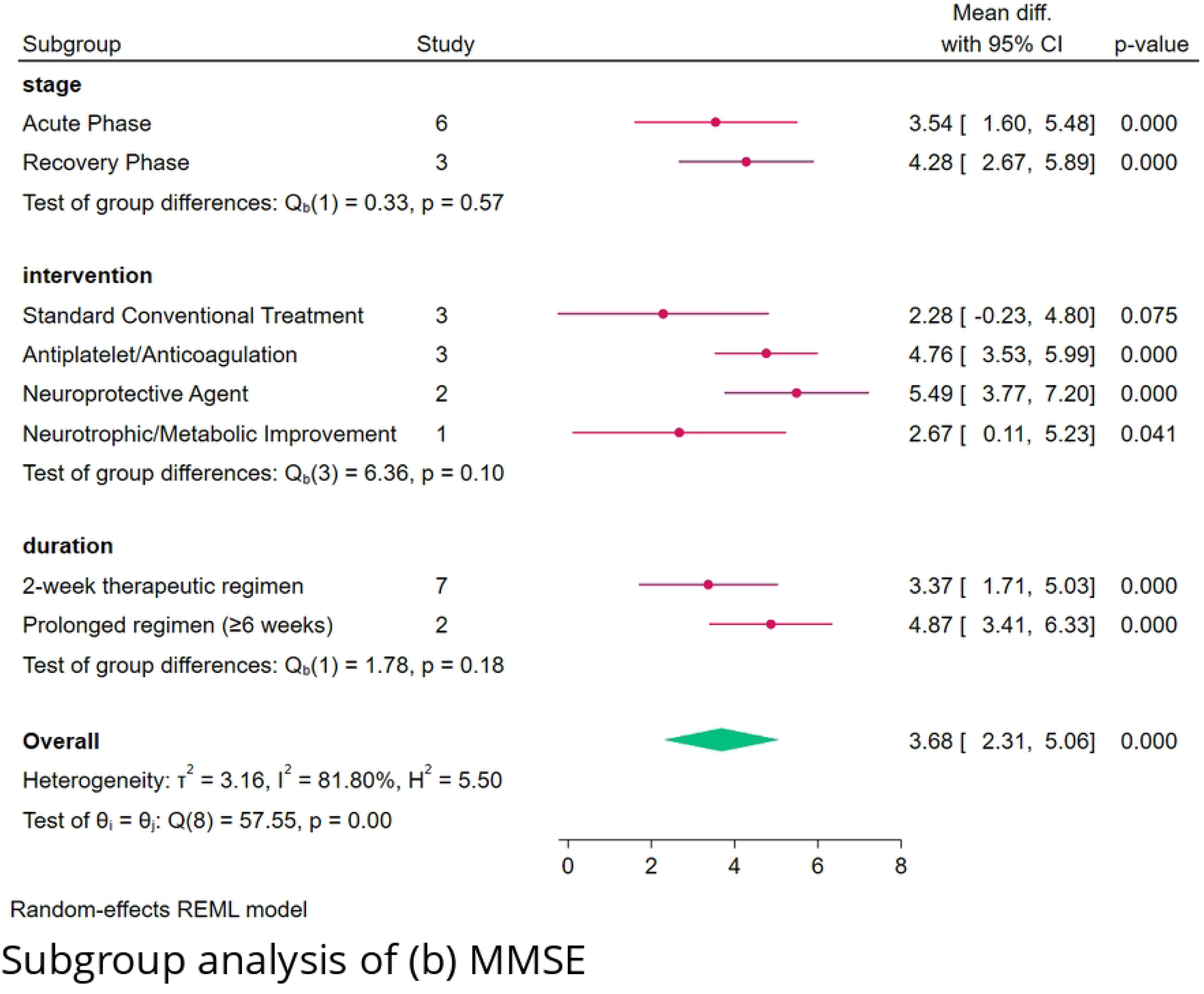
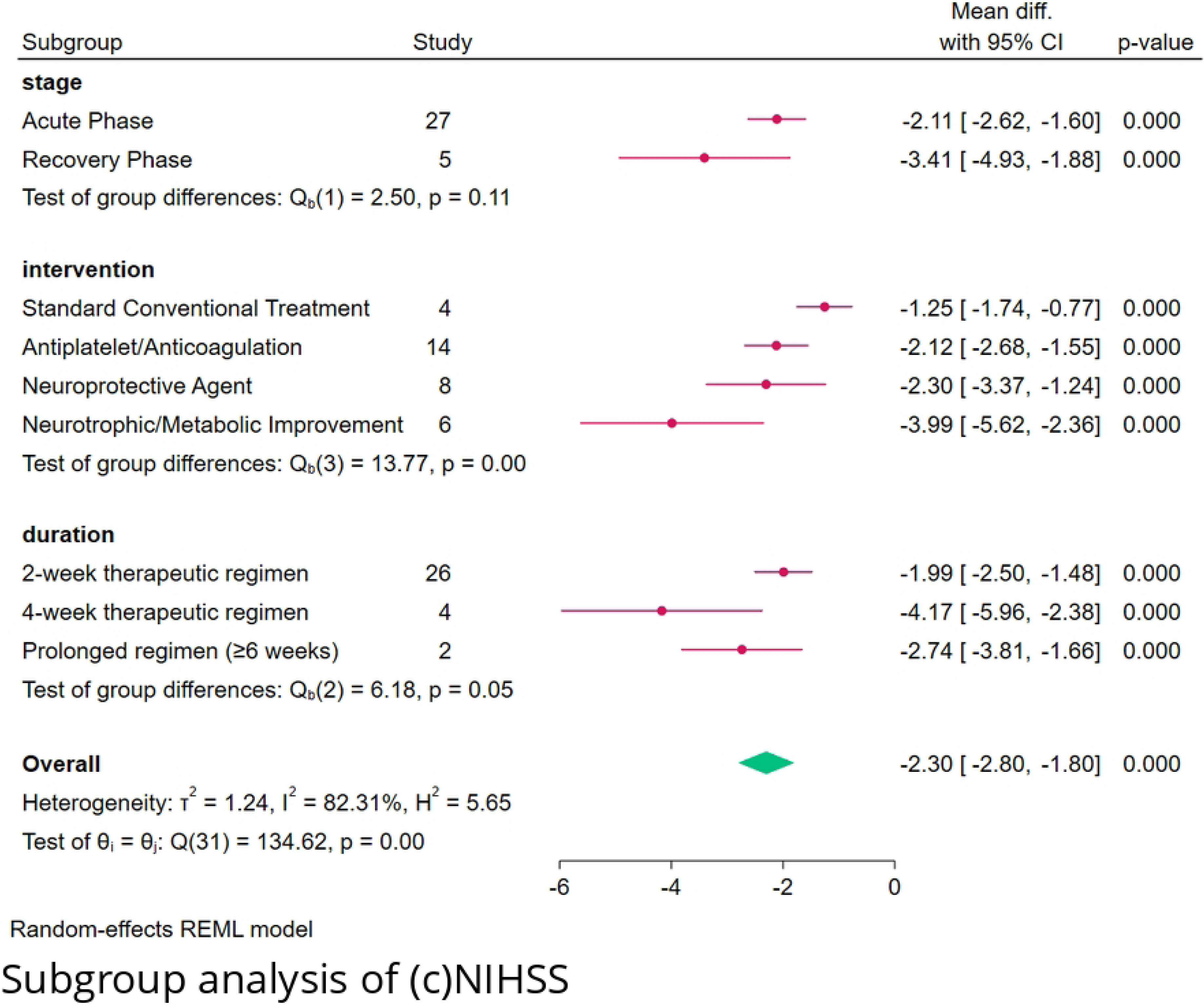
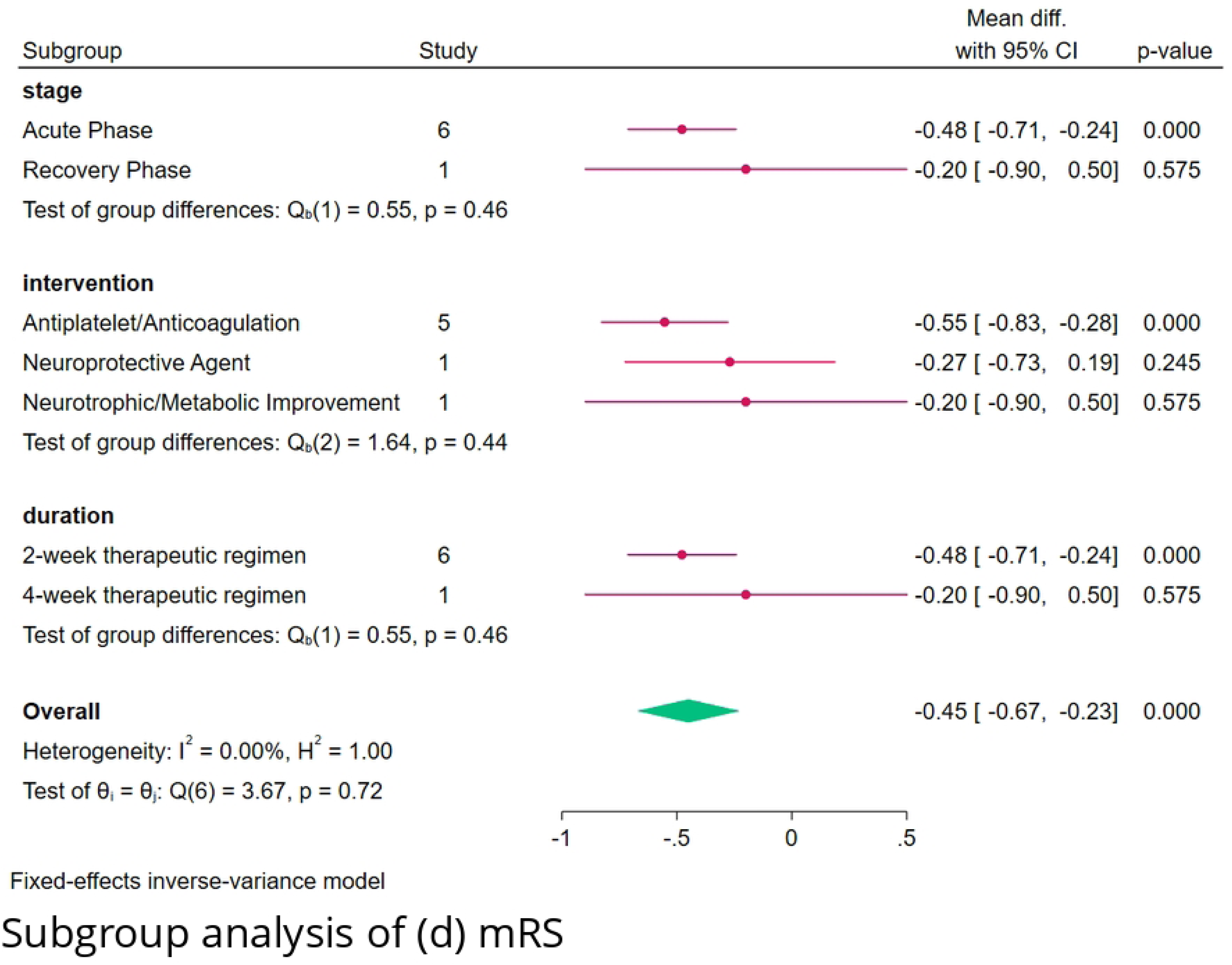
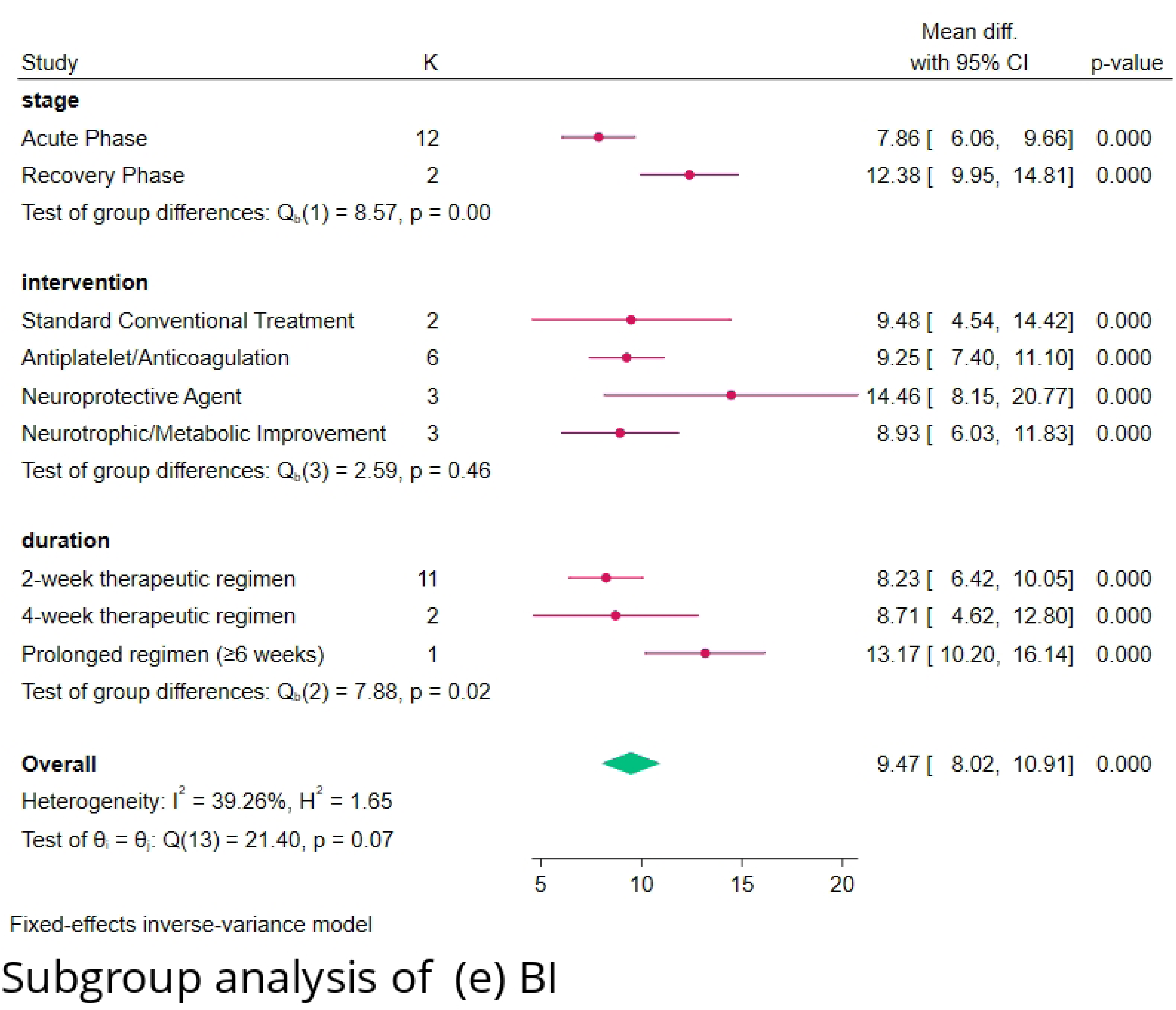
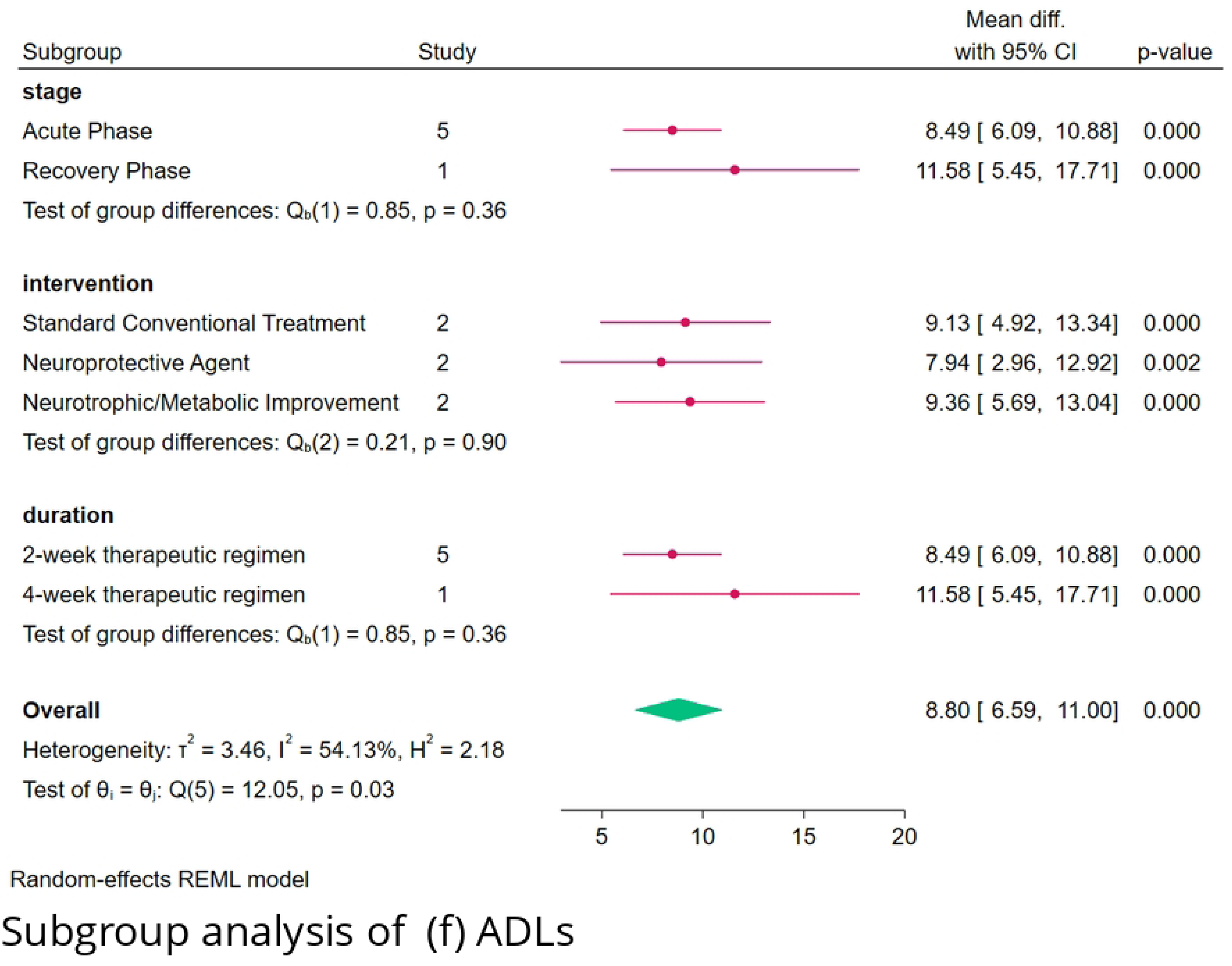

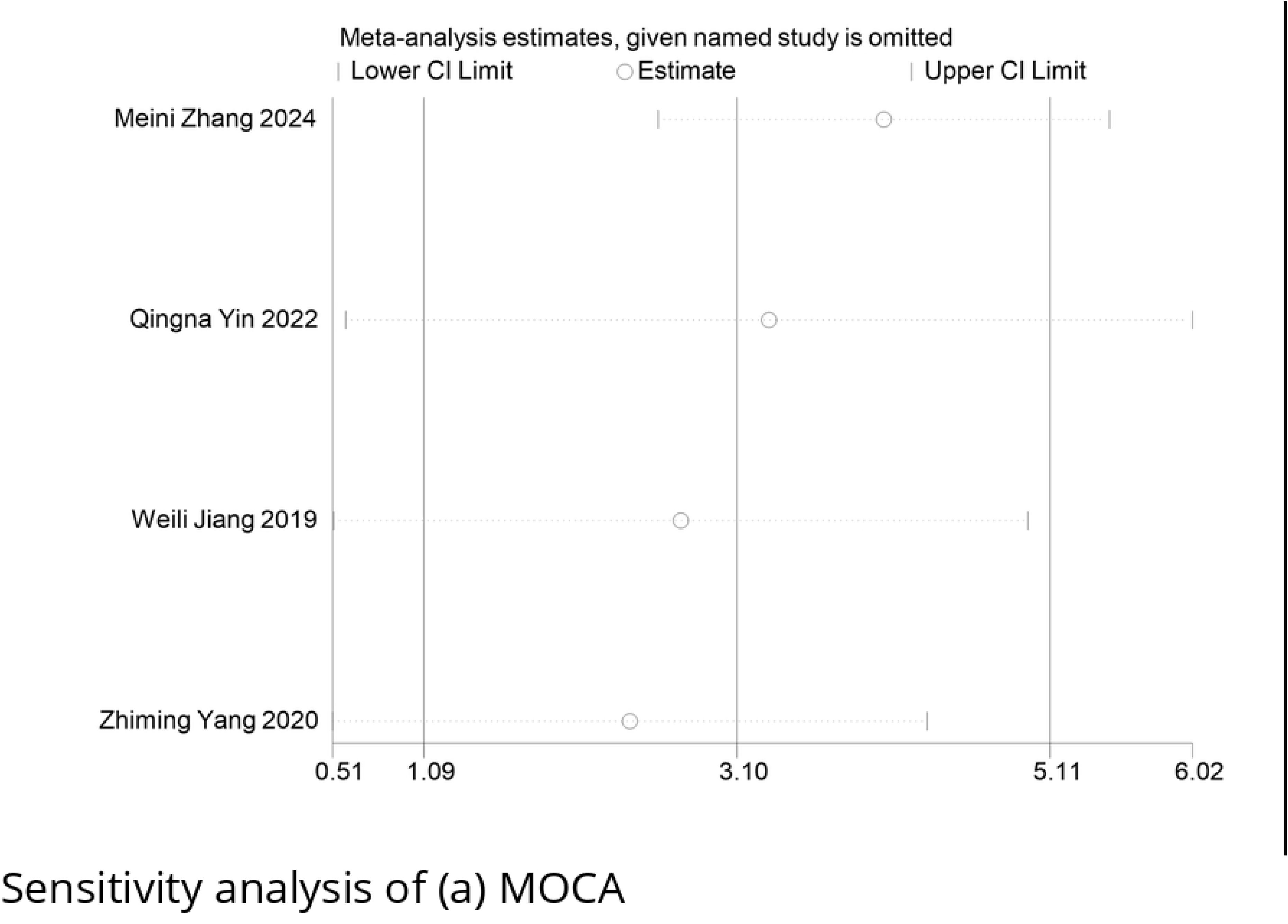
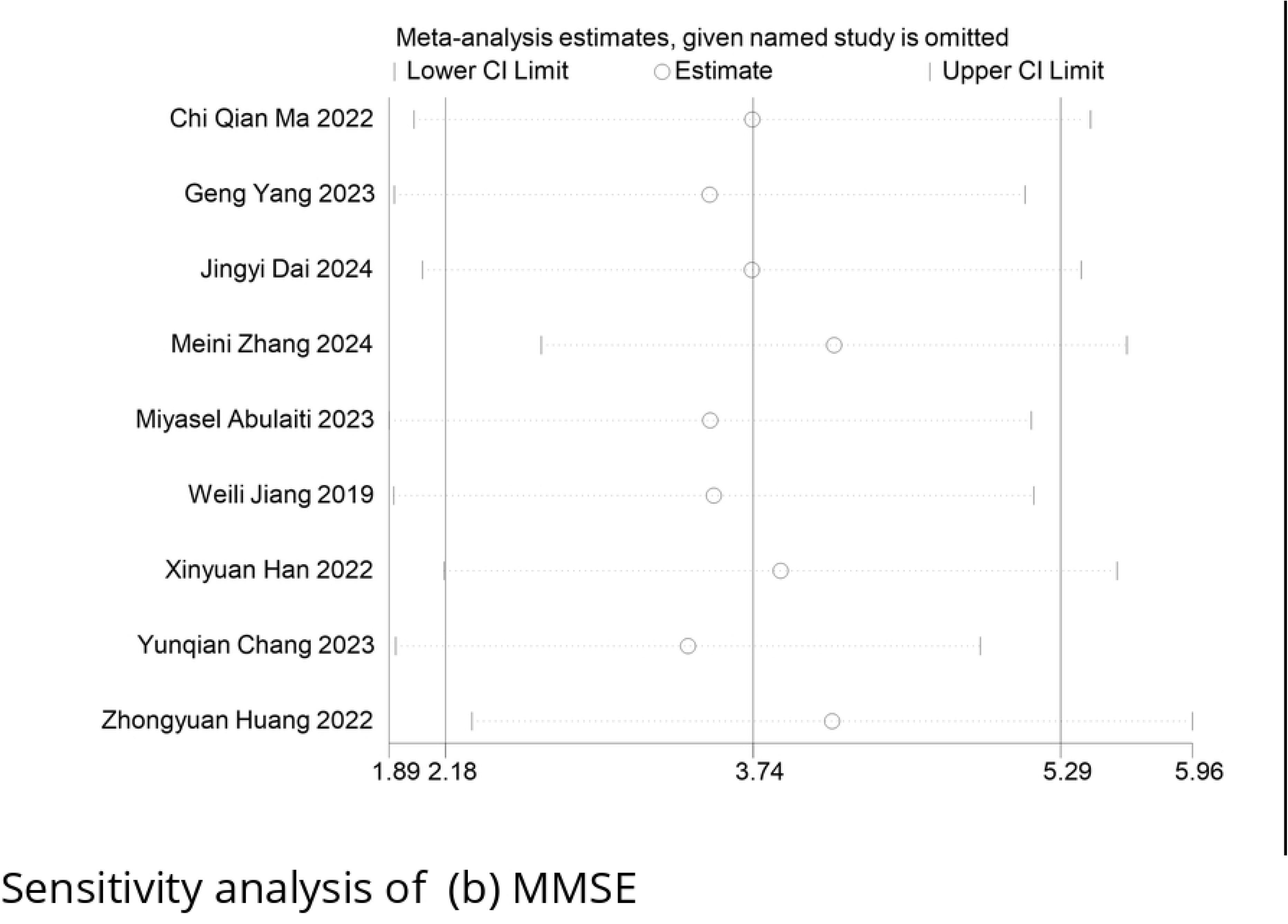
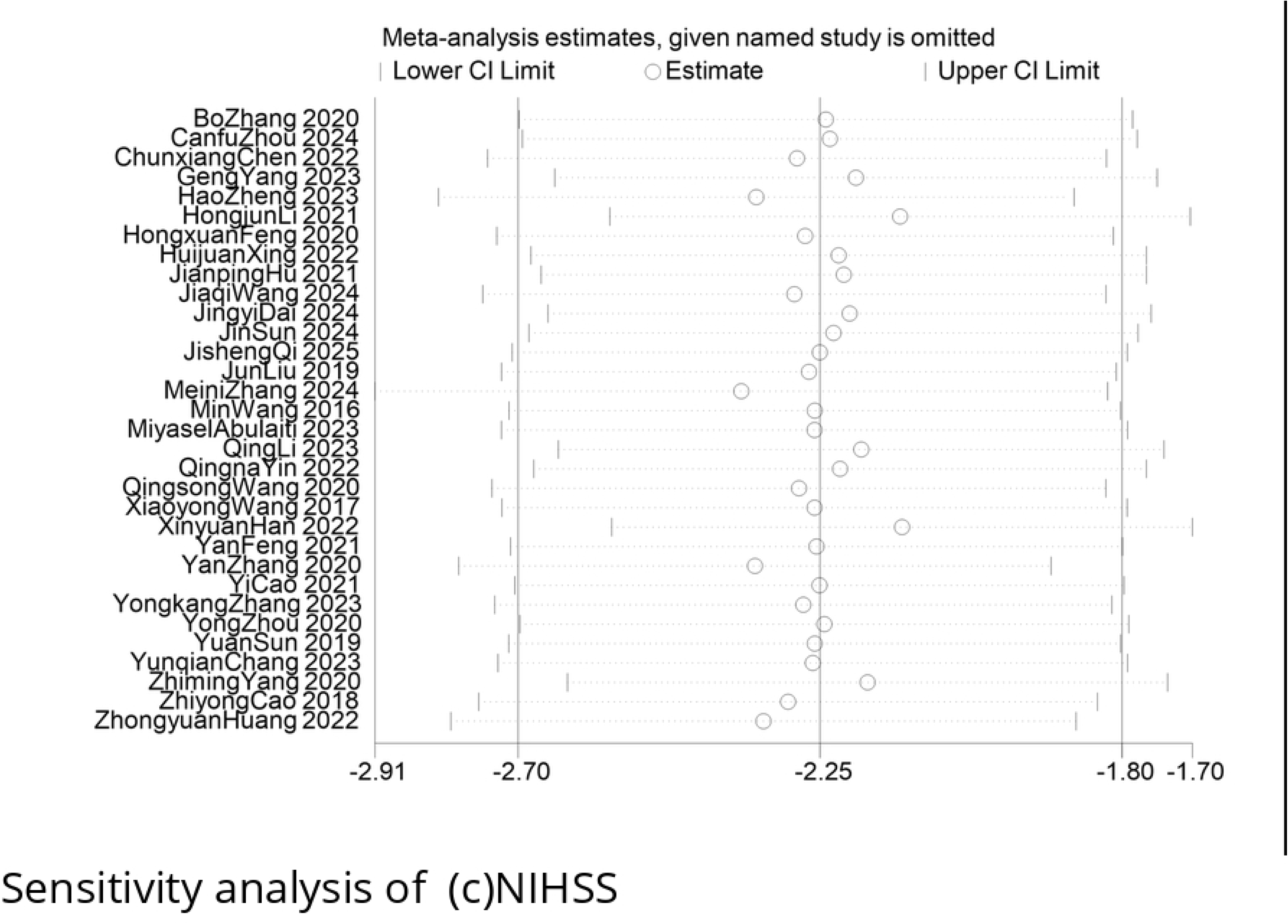
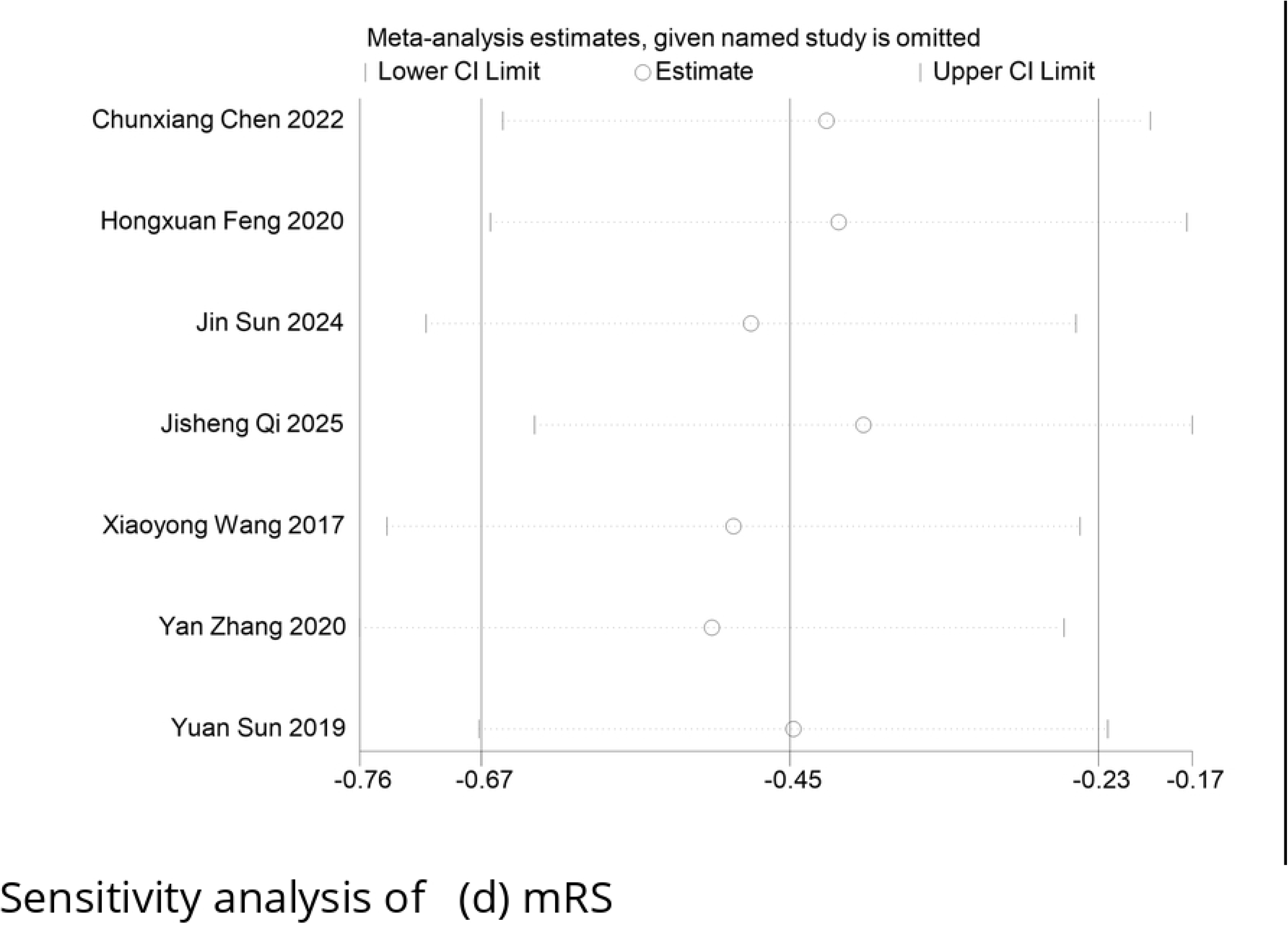
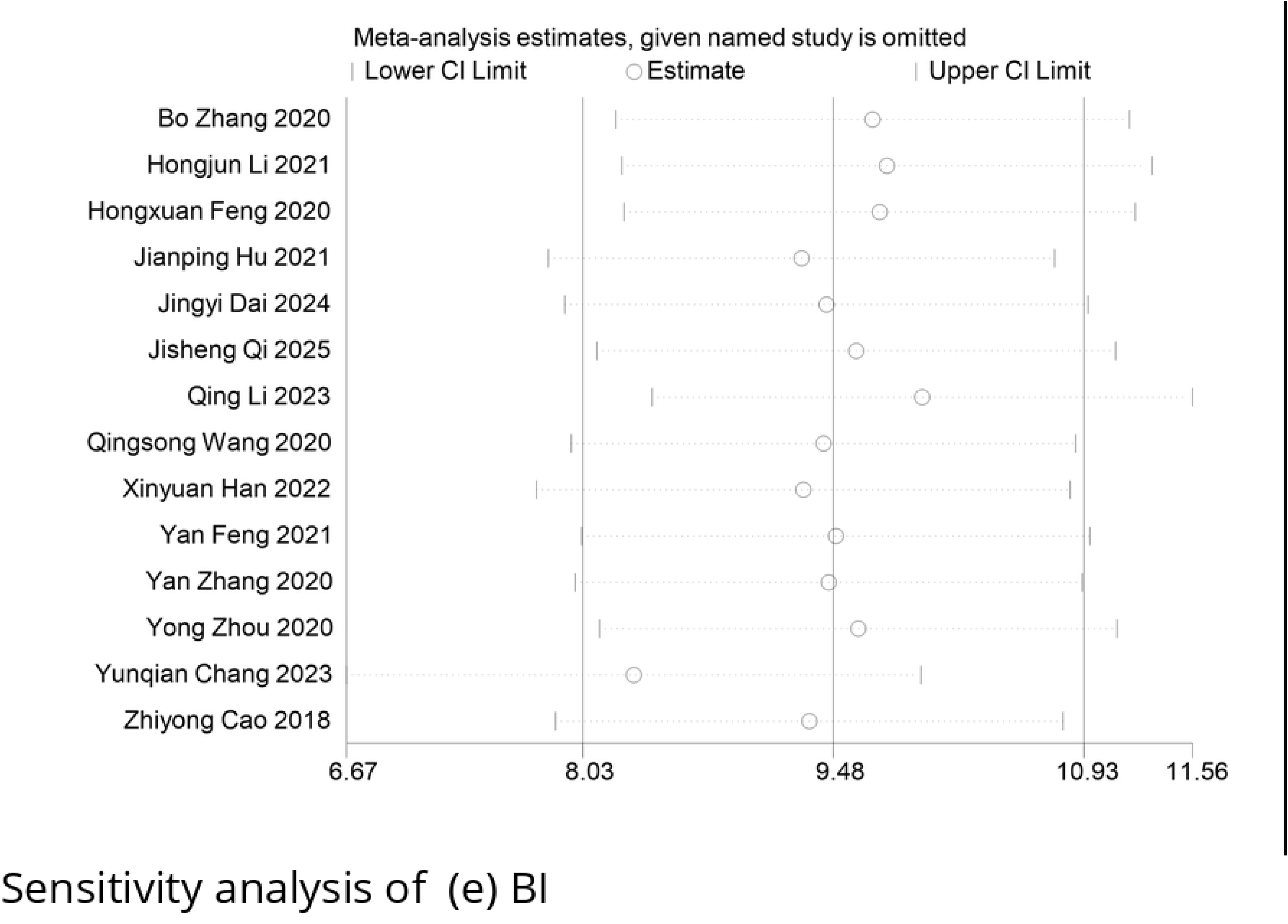
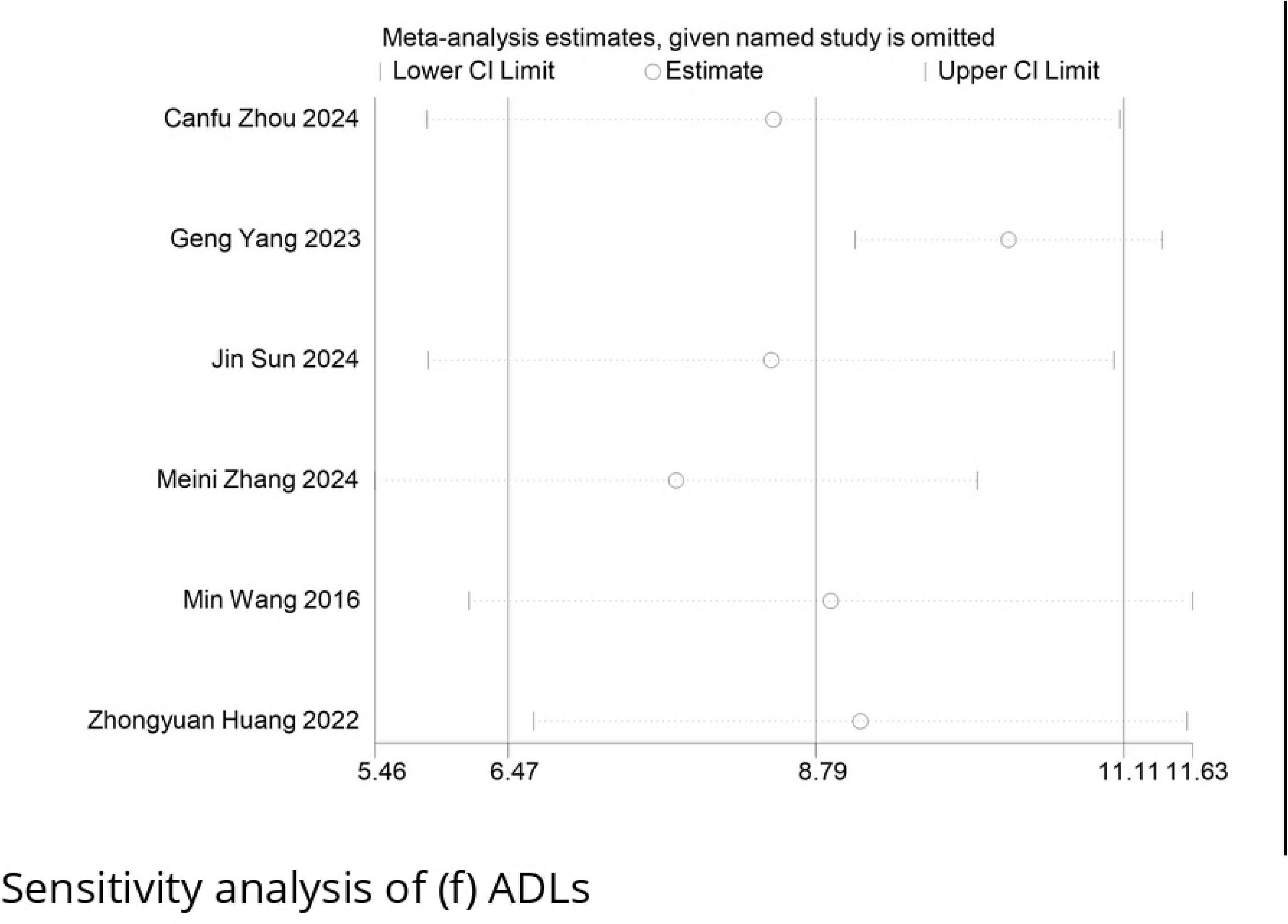

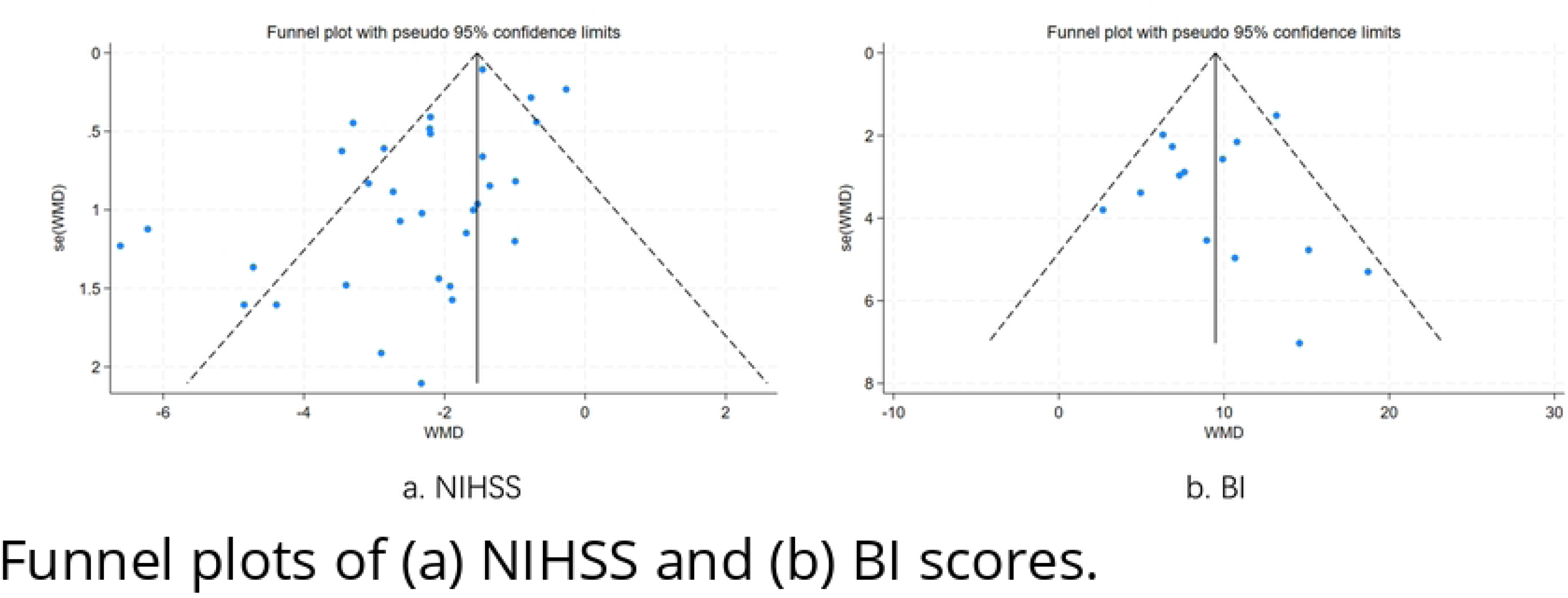

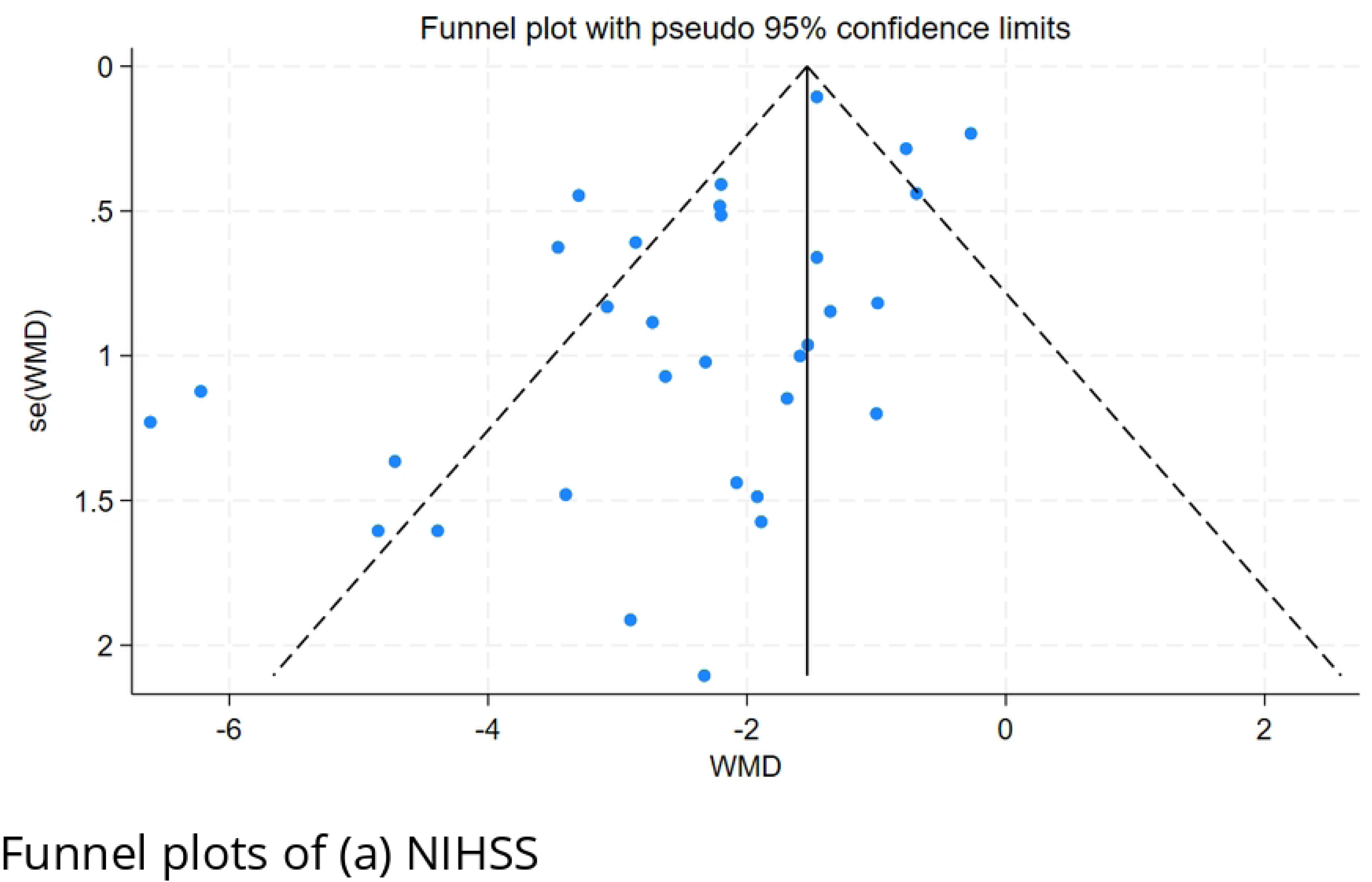
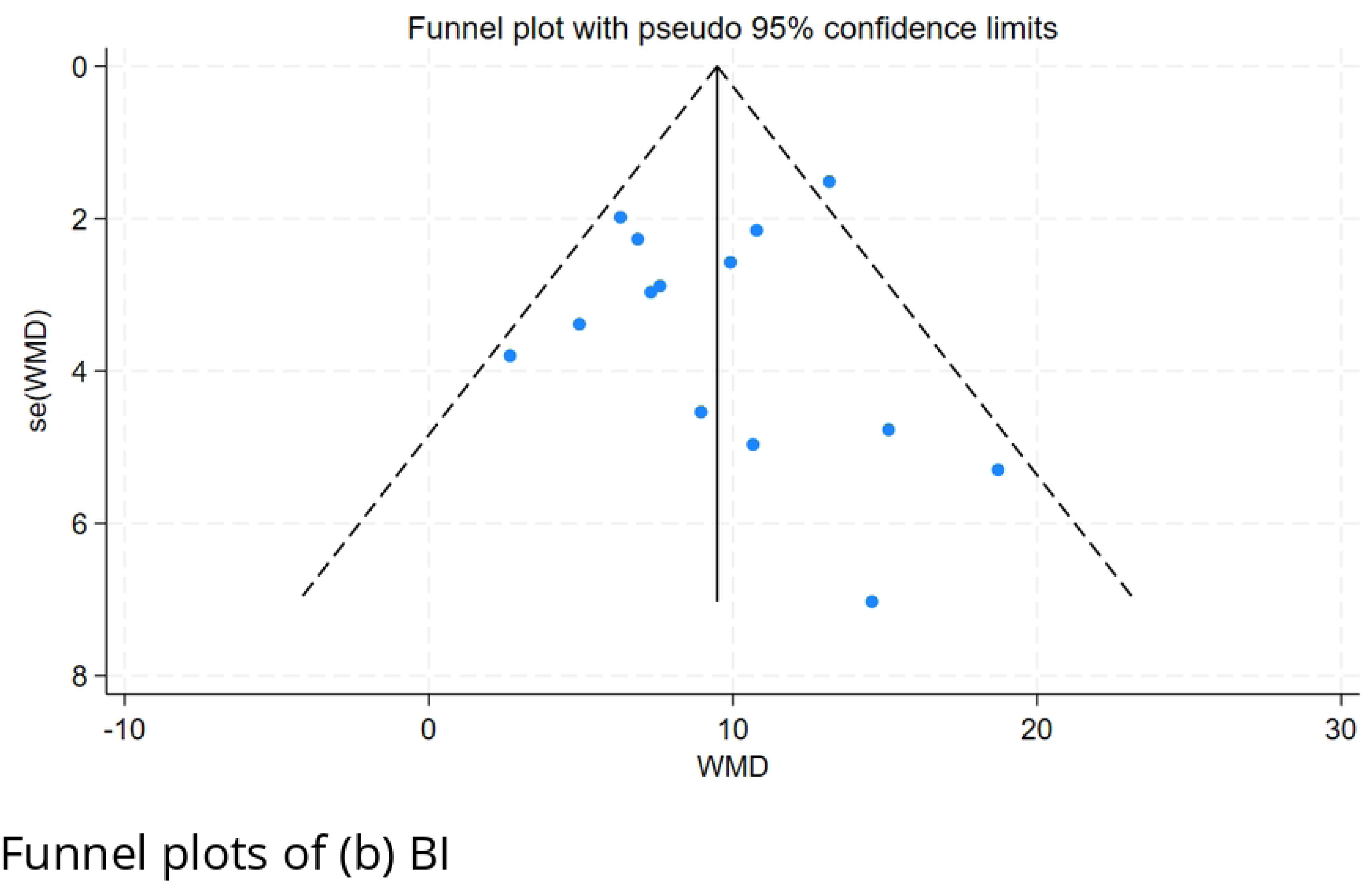

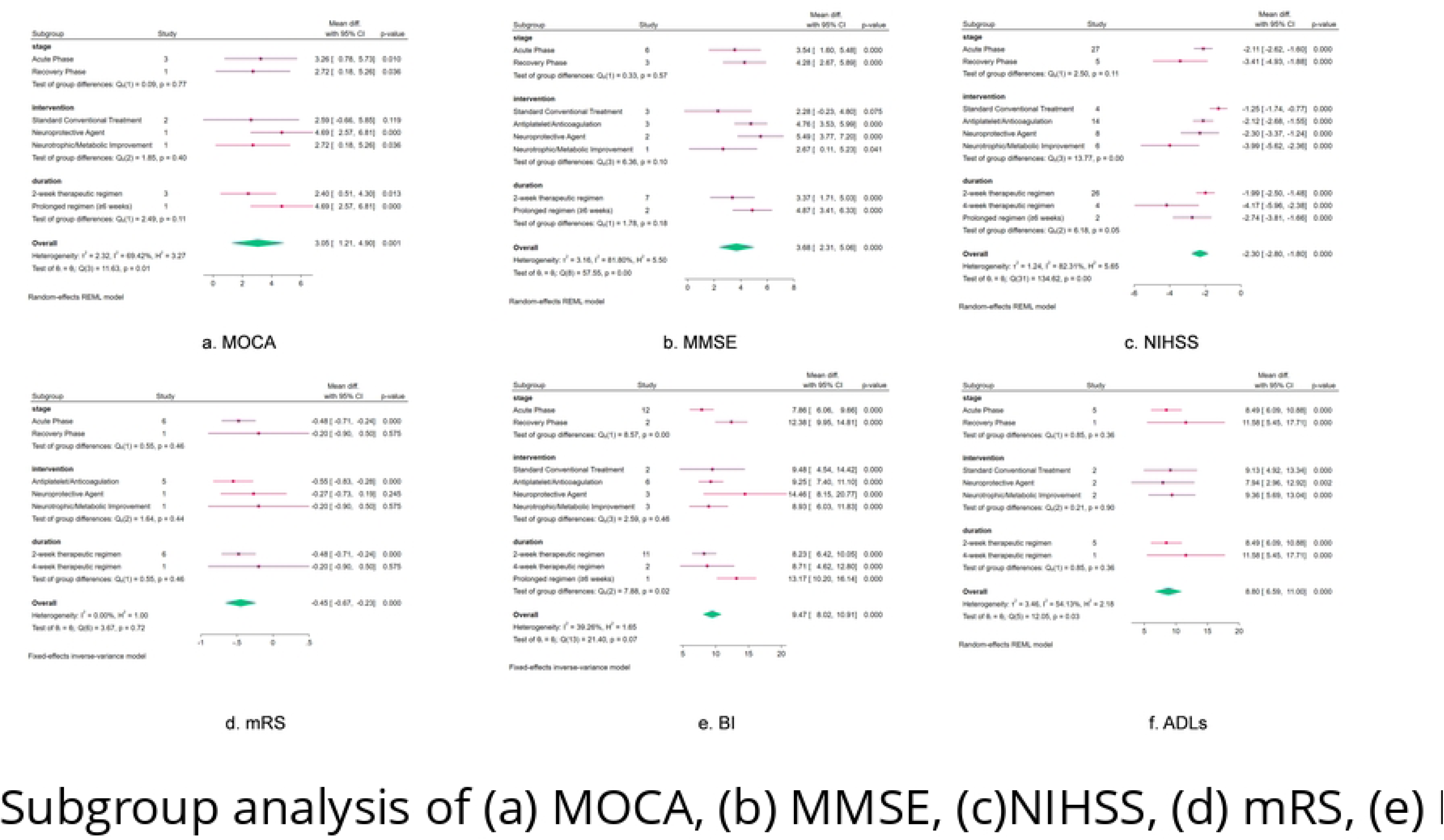

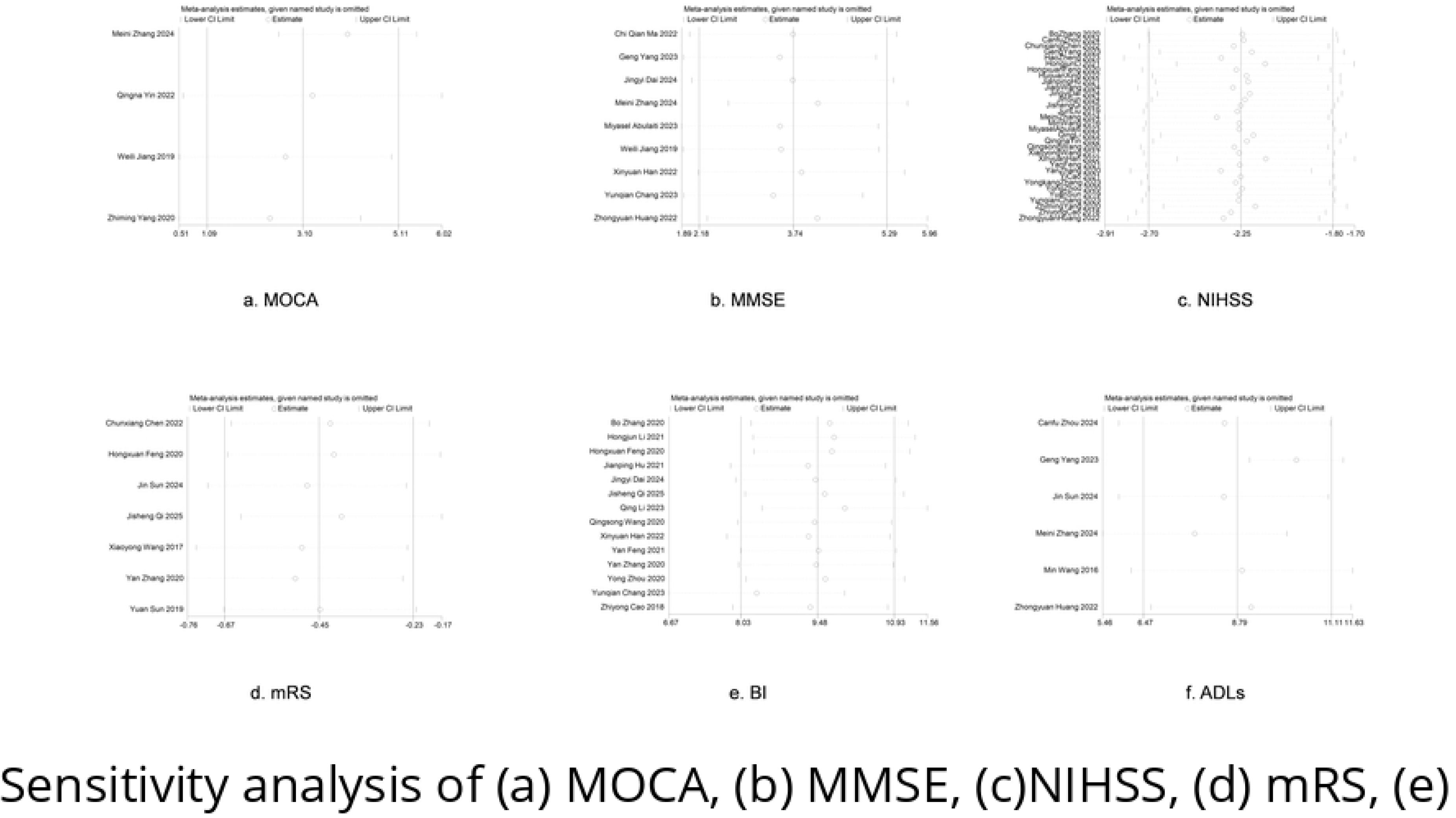

